# Non-exercise Machine Learning Models for Maximal Oxygen Uptake Prediction in National Population Surveys

**DOI:** 10.1101/2022.09.30.22280471

**Authors:** Yuntian Liu, Jeph Herrin, Chenxi Huang, Rohan Khera, Lovedeep Singh Dhingra, Weilai Dong, Bobak J. Mortazavi, Harlan M. Krumholz, Yuan Lu

## Abstract

**Background:** Maximal oxygen uptake (VO_2_ max), an indicator of cardiorespiratory fitness (CRF), requires exercise testing and, as a result, is rarely ascertained in large-scale population-based studies. Non-exercise algorithms are cost-effective methods to estimate VO_2_ max, but the existing models have limitations in generalizability and predictive power. This study aims to improve the non-exercise algorithms using machine learning (ML) methods and data from U.S. national population surveys.

**Methods:** We used the 1999-2004 data from the National Health and Nutrition Examination Survey (NHANES), in which a submaximal exercise test produced an estimate of the VO_2_max. We applied multiple supervised ML algorithms to build two models: a parsimonious model that used variables readily available in clinical practice, and an extended model that additionally included more complex variables from more Dual-Energy X-ray Absorptiometry (DEXA) and standard laboratory tests. We used Shapley additive explanation (SHAP) to interpret the new model and identify the key predictors. For comparison, existing non-exercise algorithms were applied unmodified to the testing set.

**Results:** Among the 5,668 NHANES participants included in the final study population, the mean age was 32.5 years and 49.9% were women. Light Gradient Boosting Machine (LightGBM) had the best performance across multiple types of supervised ML algorithms. Compared with the best existing non-exercise algorithms that could be applied in NHANES, the parsimonious LightGBM model (RMSE: 8.51 ml/kg/min [95% CI: 7.73 -9.33]) and the extended model (RMSE: 8.26 ml/kg/min [95% CI: 7.44 -9.09]) significantly reducing the error by 15% (P <0.01) and 12% (P<0.01 for both), respectively.

**Conclusion:** Our non-exercise ML model provides a more accurate prediction of VO_2_ max for NHANES participants than existing non-exercise algorithms.

**What is Known:**

- Although cardiorespiratory fitness is recognized as an important marker of cardiovascular health, it is not routinely measured because of the time and resources required to perform exercise tests.
- Non-exercise algorithms are cost-effective alternatives to estimate cardiorespiratory fitness, but the existing models are restricted in generalizability and predictive power.

**What the Study Adds:**

- We improve non-exercise algorithms for cardiorespiratory fitness prediction using advanced ML methods and a more comprehensive and representative data source from U.S. national population surveys.
- More health factors that are associated with cardiorespiratory fitness are newly identified.
- Nationally representative estimates for cardiorespiratory fitness in the U.S. over the recent 20 years are generated.

## INTRODUCTION

Cardiorespiratory fitness (CRF) refers to the integrated capacity of the circulatory and respiratory systems to transport oxygen from the atmosphere to the mitochondria to perform physical work.^1,2^ Low cardiorespiratory fitness is an important risk factor for cardiovascular disease, all-cause mortality, and mortality rates attributable to various cancers, especially of the breast and colon/digestive tract.^2-9^ Improvements in CRF are associated with reduced mortality risk.^10^

The gold standard measurement of CRF is a direct measurement of the maximal oxygen uptake (VO_2_max) using cardiopulmonary exercise testing (CPX), which combines conventional maximal exercise testing with ventilatory gas expired analysis.^11-14^ When the instrumentation and trained personnel to perform CPX are not available, a widely accepted alternative is to estimate the VO_2_max indirectly from heart rate response to a submaximal exercise.^11,15^ Although CRF is recognized as an important marker of cardiovascular health, population health surveys and cohort studies do not routinely measure CRF because of the time and resources required to perform either of these exercise tests.

As an alternative, many investigators have developed non-exercise-based algorithms to estimate CRF without performing a maximal or submaximal test. This approach uses information typically available in healthcare settings to provide an inexpensive and rapid way of estimating CRF.^8,9,16-28^ While these non-exercise algorithms are useful, they have several limitations. First, most of these algorithms were developed based on small population cohorts. The performance of these algorithms is poor when applied to other groups. Second, these algorithms were typically developed with only a limited number of health indicators available as candidate variables for prediction and could not consider a wider range of other factors that may be associated with CRF. Third, most of the previous non-exercise algorithms used linear models, and do not account for nonlinearities and interactions among the predictors.^29^ Addressing these limitations may improve the performance of the non-exercise algorithms and provide more accurate estimates of VO_2_max for research and clinical practice.

One promising approach for addressing the last limitation is machine learning (ML). Over the past decade, numerous ML applications have appeared in the latest clinical literature, especially for outcome prediction models.^30^ Given that the performance and utility of prediction models depend on the data source and methods used, a large and growing body of studies has sought to boost the existing clinical prediction models with the evolving ML approaches applied to a larger data set of a broader range of parameters. ^31,32^

Accordingly, we sought to improve the non-exercise algorithms for CRF prediction with ML methods and a more comprehensive and representative data source from U.S. national population surveys. Using data from the National Health and Nutrition Examination Survey (NHANES) and state-of-the-art ML algorithms, we aimed to overcome the limitations of the existing non-exercise algorithms and develop an approach to predict VO2max more accurately without the need for an exercise test.

## METHODS

This cross-sectional study received an exemption for review from the Institutional Review Board at Yale University because NHANES data are publicly available and de-identified. The study was reported following the Strengthening the Reporting of Observational Studies in Epidemiology (STROBE) reporting guidelines.^33^

### Data source

Data from NHANES were used for model development, validation, and prediction in this study. The NHANES is a series of cross-sectional, weighted, multi-stage sampled surveys that provide nationally representative estimates of the non-institutionalized US population. Survey participants received in-home interviews, followed by standardized physical examinations conducted in mobile examination centers, and laboratory tests using blood and urine specimens provided by participants during the physical examination. Since 1999, NHANES has become a continuous program conducted in 2-year cycles. However, due to the coronavirus disease 2019 (COVID-19) pandemic, data collection for the latest NHANES 2019-2020 cycle was suspended in March 2020 and the data collected from 2019 to March 2020 were combined with data from the NHANES 2017-2018 cycle to form a nationally representative sample of NHANES 2017-March 2020 pre-pandemic data.^34^

### Study population

We used data from 3 cycles conducted from 1999-2000 through 2003-2004 for model development and validation because CRF was measured only for that period in NHANES. Participants for CRF measurements were selected based on age (age eligibility for CRF measurement in NHANES: 12-49 years), medical conditions, medications, physical limitations, heart rate and blood pressure. The screening was done before the treadmill test using questions in the household interview, questions administered by the physician in the NHANES Mobile Examination Center (MEC), and measurements of heart rate and blood pressure. The list of exclusion criteria used for the CRF component in NHANES was attached in the supplemental material (**Supplemental Table S1**). We further limited our study population to participants aged 16-49 years because of the age restriction in the NHANES physical activity data section.

### Study outcome

Our primary outcome of interest was VO_2_max. NHANES’s protocol for measuring VO_2_max (ml/kg/min) is the submaximal exercise test. Based on gender, age, body mass index, and self-reported level of physical activity, participants were assigned to one of eight treadmill test protocols. Heart rate was monitored continuously using an automated monitor with four electrodes connected to the thorax and abdomen of the participant and was recorded at the end of the warm-up, each exercise stage, and each minute of recovery. Then VO_2_max is then estimated by extrapolation using measured heart rate responses to known levels of exercise workloads in the CRF exam, assuming the relation between heart rate and oxygen consumption is linear during exercise.^11^ Detailed descriptions of the protocol are provided in the NHANES Cardiovascular Fitness Procedure Manual.^35^

### Candidate variables

To develop a data-driven model, we considered a variety of predictors available in NHANES, including variables from home interview, physical examination, and laboratory data section. Variables used in this study were selected based on potential associations with overall health status, cardiovascular health, respiratory system, and metabolic systems, according to a systematic review.^2^ Then selected variables were examined for their continuities, to see if they were consistently available or at least can be estimated by other consistently available variables throughout the specified NHANES data collection period (1999-March 2020). This was important for the model’s generalizability and sustainability in NHANES. The exception was the daily alcohol intake (not available in 2019-March 2020) and Dual-Energy X-ray Absorptiometry (DEXA) data (not available in 2007-2010 and 2019-March 2020). DEXA data in NHANES contains information about body composition like the fat percentage of different body parts, which is important to the prediction of CRF.^4,16,24,25,28^ Finally, a total of 49 variables from different data sections were selected in the final feature set. (**Supplemental Table S4**). We applied a log-transformation to the reported weekly moderate-intensity physical activity time to account for its skewed distribution. All the 49 selected variables in the final study population had a missing rate of less than 10%, except for albuminuria (10.4%). We explained detailed methods to address missing data in the following statistical analysis section.

### Existing non-exercise algorithms

We exhaustively searched for non-exercise algorithms that were developed in previous studies^16,18,23-25,28^ or presented in systematic reviews^2,36^ as baseline models for both validation and comparison in this study. The existing algorithms were selected based on the following two criteria: (1) The non-exercise algorithm targeted a relatively large and general population in the U.S. (2) The non-exercise algorithms used the variables that are available or can be approximated in NHANES, and the algorithm itself can be applied in the context of NHANES. These selected non-exercise algorithms were applied and evaluated as they were, i.e., using their original parameters or coefficients. Detailed information on the final algorithms considered is reported in **Table 1**.

**Table 1.**
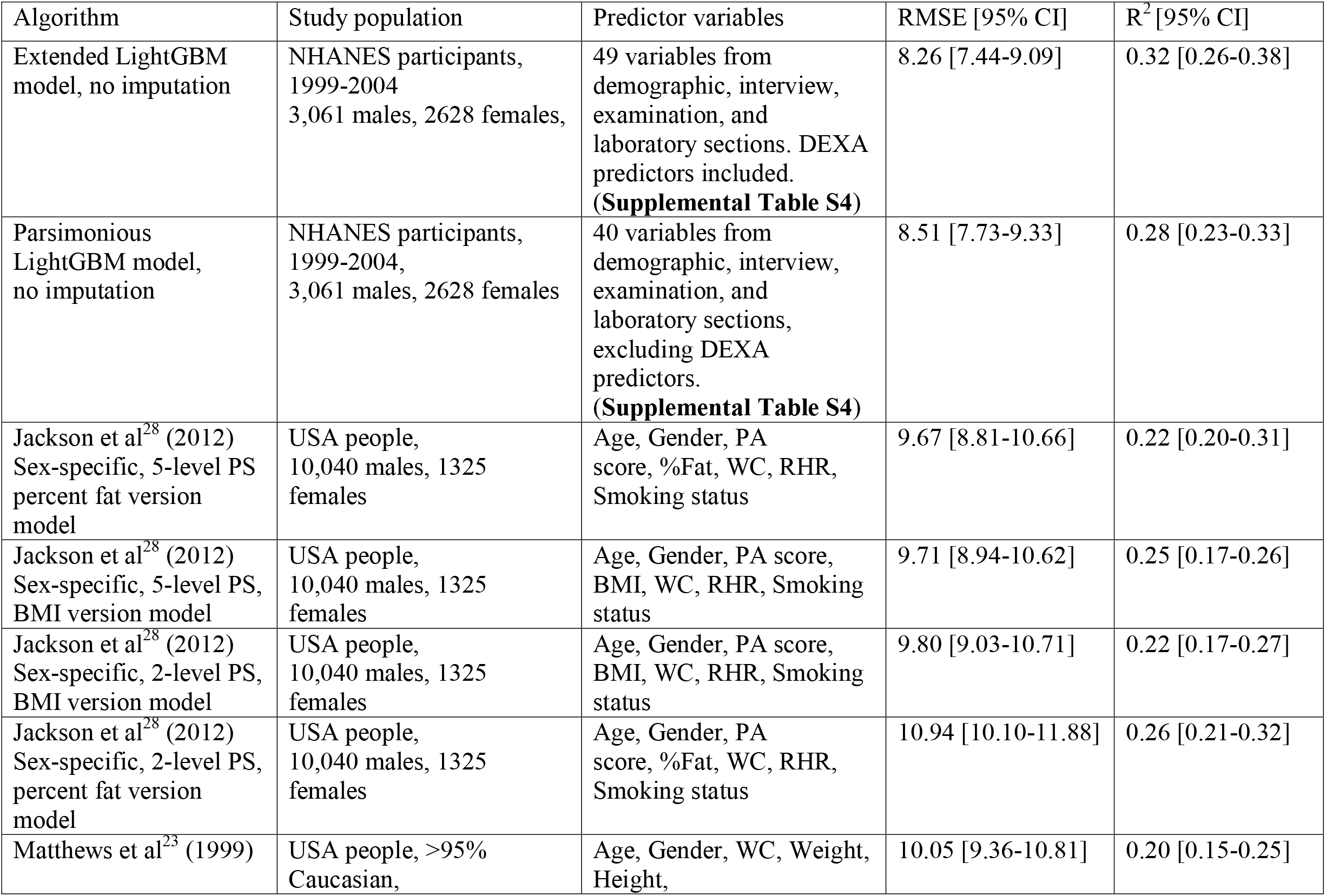
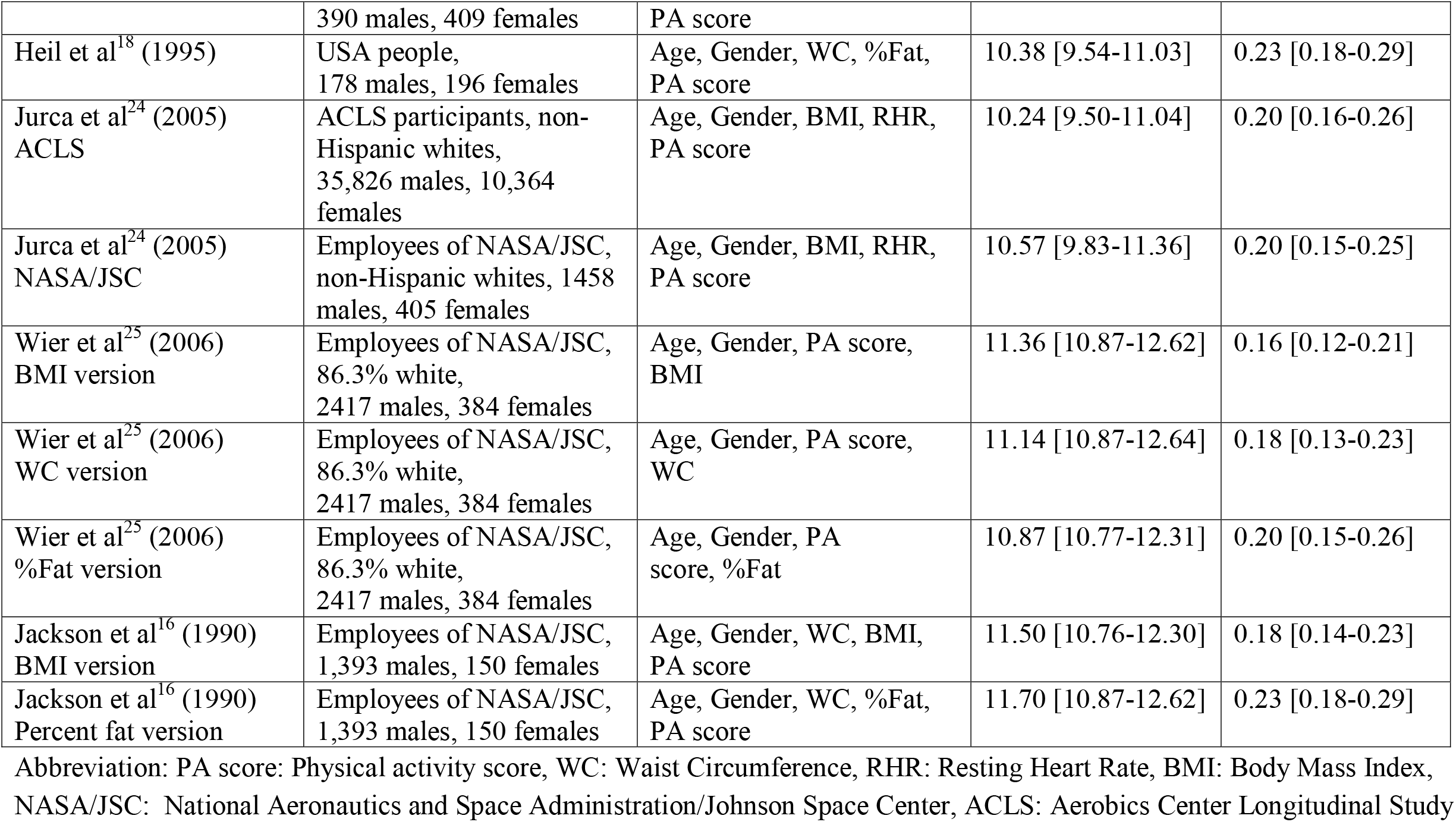
Comparison of newly developed optimal ML model and the existing non-exercise algorithms

| Algorithm | Study population | Predictor variables | RMSE [95% CI] | R<sup>2</sup> [95% CI] |
| --- | --- | --- | --- | --- |
| Extended LightGBM model, no imputation | NHANES participants, 1999-2004<br>3,061 males, 2628 females, | 49 variables from demographic, interview, examination, and laboratory sections. DEXA predictors included.<br>**(Supplemental Table S4)** | 8.26 [7.44-9.09] | 0.32 [0.26-0.38] |
| Parsimonious LightGBM model, no imputation | NHANES participants, 1999-2004,<br>3,061 males, 2628 females | 40 variables from demographic, interview, examination, and laboratory sections, excluding DEXA predictors.<br>**(Supplemental Table S4)** | 8.51 [7.73-9.33] | 0.28 [0.23-0.33] |
| Jackson et al<sup>28</sup> (2012)<br>Sex-specific, 5-level PS percent fat version model | USA people, 10,040 males, 1325 females | Age, Gender, PA score, %Fat, WC, RHR, Smoking status | 9.67 [8.81-10.66] | 0.22 [0.20-0.31] |
| Jackson et al<sup>28</sup> (2012)<br>Sex-specific, 5-level PS, BMI version model | USA people, 10,040 males, 1325 females | Age, Gender, PA score, BMI, WC, RHR, Smoking status | 9.71 [8.94-10.62] | 0.25 [0.17-0.26] |
| Jackson et al<sup>28</sup> (2012)<br>Sex-specific, 2-level PS, BMI version model | USA people, 10,040 males, 1325 females | Age, Gender, PA score, BMI, WC, RHR, Smoking status | 9.80 [9.03-10.71] | 0.22 [0.17-0.27] |
| Jackson et al<sup>28</sup> (2012)<br>Sex-specific, 2-level PS, percent fat version model | USA people, 10,040 males, 1325 females | Age, Gender, PA score, %Fat, WC, RHR, Smoking status | 10.94 [10.10-11.88] | 0.26 [0.21-0.32] |
| Matthews et al<sup>23</sup> (1999) | USA people, >95% Caucasian, | Age, Gender, WC, Weight, Height, | 10.05 [9.36-10.81] | 0.20 [0.15-0.25] |
|  | 390 males, 409 females | PA score |  |  |
| Heil et al<sup>18</sup> (1995) | USA people,<br>178 males, 196 females | Age, Gender, WC, %Fat,<br>PA score | 10.38 [9.54-11.03] | 0.23 [0.18-0.29] |
| Jurca et al<sup>24</sup> (2005)<br>ACLS | ACLS participants, non-Hispanic whites,<br>35,826 males, 10,364<br>females | Age, Gender, BMI, RHR,<br>PA score | 10.24 [9.50-11.04] | 0.20 [0.16-0.26] |
| Jurca et al<sup>24</sup> (2005)<br>NASA/JSC | Employees of NASA/JSC,<br>non-Hispanic whites, 1458<br>males, 405 females | Age, Gender, BMI, RHR,<br>PA score | 10.57 [9.83-11.36] | 0.20 [0.15-0.25] |
| Wier et al<sup>25</sup> (2006)<br>BMI version | Employees of NASA/JSC,<br>86.3% white,<br>2417 males, 384 females | Age, Gender, PA score,<br>BMI | 11.36 [10.87-12.62] | 0.16 [0.12-0.21] |
| Wier et al<sup>25</sup> (2006)<br>WC version | Employees of NASA/JSC,<br>86.3% white,<br>2417 males, 384 females | Age, Gender, PA score,<br>WC | 11.14 [10.87-12.64] | 0.18 [0.13-0.23] |
| Wier et al<sup>25</sup> (2006)<br>%Fat version | Employees of NASA/JSC,<br>86.3% white,<br>2417 males, 384 females | Age, Gender, PA<br>score, %Fat | 10.87 [10.77-12.31] | 0.20 [0.15-0.26] |
| Jackson et al<sup>16</sup> (1990)<br>BMI version | Employees of NASA/JSC,<br>1,393 males, 150 females | Age, Gender, WC, BMI,<br>PA score | 11.50 [10.76-12.30] | 0.18 [0.14-0.23] |
| Jackson et al<sup>16</sup> (1990)<br>Percent fat version | Employees of NASA/JSC,<br>1,393 males, 150 females | Age, Gender, WC, %Fat,<br>PA score | 11.70 [10.87-12.62] | 0.23 [0.18-0.29] |

Abbreviation: PA score: Physical activity score, WC: Waist Circumference, RHR: Resting Heart Rate, BMI: Body Mass Index, NASA/JSC: National Aeronautics and Space Administration/Johnson Space Center, ACLS: Aerobics Center Longitudinal Study

### Machine learning algorithms

We used multiple types of supervised machine learning algorithms for the modelling of VO_2_max using the candidate variables. The algorithms were K-Nearest Neighbors (KNN),^37^ Least Absolute Shrinkage and Selection Operator (LASSO),^38^ Support Vector Regression (SVR),^39^ Random Forest (RF),^40^ and Gradient Boosting decision tree^41^ (GBDT) family which included Extreme Gradient Boosting (XGBoost) and Light Gradient Boosting Machine (LightGBM).^42,43^ The detailed descriptions of these algorithms are listed in **Supplemental Table S3**.

### Model development

We built two types of models, the parsimonious model, and the extended model, according to the range of selected feature sets. The parsimonious model was trained on only variables that are easily available in normal clinical settings, mostly from the demographics, interviews, and regular physical examination. The extended model was fitted using all the selected candidate variables, including standard lab test results and DEXA data. We developed two types of models because the parsimonious model may be more applicable in common practice, while the extended model may have better predictive power and could shed light on the unexplored associations between CRF and other potential factors.

We split the final study population (N=5668) into a training set (80%) and a validation set (20%). Given the continuous outcome VO_2_max, each ML model was trained for a regression task using root mean square error (RMSE) as the loss function. Optimal hyperparameters (parameters whose values are used to control the learning process) were tuned using a 100 times random search with 5-fold cross-validation on the training set. Specifically, with each set of hyperparameters chosen at random, the training set was first randomly partitioned into 5 equal subsets (folds) and each fold was left out to be the hold-out fold in turn. For each hold-out fold, we repeated the process of training the ML algorithm with the specific set of hyperparameters on the remaining four folds and assessing the performance on this hold-out set. The optimal set of hyperparameters was selected according to a minimum averaged RMSE over the 5 assessments to the 5 hold-out folds. For each version of the different ML algorithms, a final model was obtained by retraining the algorithm on the entire training set with the optimal hyperparameters, and the performance of this final model was assessed on the validation set.

### Model evaluation and model application

We evaluated each final model’s performance based on the Root-Mean-Square Error (RMSE) and the coefficient of determination (R^2^) with the 95% confidence intervals determined using bootstrapping with 1000 replicates from the validation set. Then we applied our optimal ML prediction models to NHANES cycles after 2005, where CRF was not measured. Specifically, we used the parsimonious model to predict VO_2_max for each eligible NHANES subject (**Supplemental Table S2**) from 2005 through 2020 and applied our extended model to eligible participants from 2005-2006 and 2011-2018 (DEXA data were not available for 2007-2010 and 2019-March 2020). Finally, we respectively generated the corresponding predicted nationally representative estimates for the different periods and compared them to the exercise test estimates from 1999-2004.

### Feature importance

We assessed feature importance using SHAP^44,45^ (Shapley Additive Explanation) values to identify the relative contribution of a feature to the final prediction. SHAP values are based on Shapley values which are derived from conditional game theory, attempting to fairly distribute the prediction among all the features.^46^ SHAP generalizes the Shapley values to predictive modeling and quantifies the contribution that each feature brings to the prediction made by the model.

### Statistical Analysis

We assigned equal weight to each NHANES participant in our final study population when developing the models, thus we did not consider the sampling structure of survey data in the phase of model development. To deal with the missingness, we adopted two strategies according to the algorithms we used. For LASSO, KNN, SVR, and RF, we imputed the missing data using multivariate imputation by chained equation.^47^ For XGBoost and LightGBM in the GBDT family, we took advantage of their built-in functionality, which ignores missing values during a split when a decision tree grows, then allocates them to whichever side reduces the loss/error the most, to address missingness without extra missing value imputation. We also applied the GBDTs models to the imputed data to compare the two strategies of handling missing values in this study. Categorical features were converted into a binary format using the 1-hot encoding (ie, variables with k categories were transformed into k binary indicator variables).

While generating nationally representative estimates, we incorporated the corresponding strata and weights for the estimation as per NHANES requirement. Participants’ weights were pooled and divided by the number of years studied, following the NHANES guidance. We reported descriptive statistics of participant characteristics, in the nationally representative form, as the mean (SD) or mean (range) for continuous variables and as the percentage for categorical variables, as appropriate. All statistical tests were two-sided, with a level of significance of 0.05. Data analysis was performed using R (version 4.0.1) and Python (version 3.8.5).

## RESULTS

### Baseline Characteristics

From NHANES 1999-2004, we included 5,668 participants in our final study population for model development. Among the national representative estimates of the baseline characteristics, the mean (SD) age was 32.5 (10.0) years; 49.9% were female, 69.5% were White, 15.4% were Hispanic, 10.6% were Black, and 5% belonged to the other race and ethnicity. Additionally, 21.8% of the study population lacked coverage of health insurance, 30.7% were in a low family income level, 18.0% were unemployed or not in the labor force, and 55.7% had a degree greater than high school. The observed values of VO2 max had a mean (range) of 40.1 (18.2-132.1).

### Model comparison

Among the existing non-exercise algorithms assessed in the validation set, Jackson’s sex-specific algorithms^28^ in general had the best performance: the version using a 5-level physical activity score and BMI provided the best combination of RMSE (9.71 [95% CI: 8.94-10.62]) and R^2^ (0.25 [95% CI: 0.17-0.26]). Jackson’s sex-specific algorithms were followed by the models from Matthews^23^ and Jurca^24^, whose RMSEs were slightly over 10. Other methods had relatively poor performances, with RMSEs greater than 11. Detailed information for the existing non-exercise models is listed in **Table 1**.

As for the newly developed parsimonious ML models, the LightGBM model with the algorithm’s default solution for the missingness performed the best with an RMSE of 8.51 [95% CI: 7.73-9.33] and an R^2^ of 0.28 [95% CI: 0.23-0.33] on the validation set. Among the extended ML models that include all candidate variables, the LightGBM model with the algorithm’s default solution for the missingness also outperformed the others with an RMSE of 8.26 [95% CI: 7.44-9.09] and an R^2^ of 0.32 [95%CI: 0.26-0.38] on the validation set (**Supplemental Figure S5**). The parsimonious and the extended LightGBM models significantly reduced the error by 12% (P <0.001) and 15% (P<0.001) compared with the best existing non-exercise model verified in this study, respectively (**Figure 2**). Although the extended LightGBM model performed significantly better than the parsimonious one (P <0.05), the agreement between these two models was strong since according to the Bland-Altman plot (**Supplemental Figure S6**), there was no significant systematic difference. We also investigated the distributions of the differences between the observations and the predictions provided by either the optimal ML models or the existing non-exercise algorithms. Based on the density curves of those distributions (**Supplemental Figure S7**), among the optimal ML models which in general had smaller biases than the existing algorithms, there were no systematic or consistent differences in their biases. Thus, the parsimonious model, though not as accurate as the extended model, can be a reliable alternative in most cases where the extended model is not applicable. Other newly developed ML models in this study also generally outperformed the existing non-exercise algorithms (**Table1, Figure 2, Supplemental Figure S5**).

**Figure 1.**
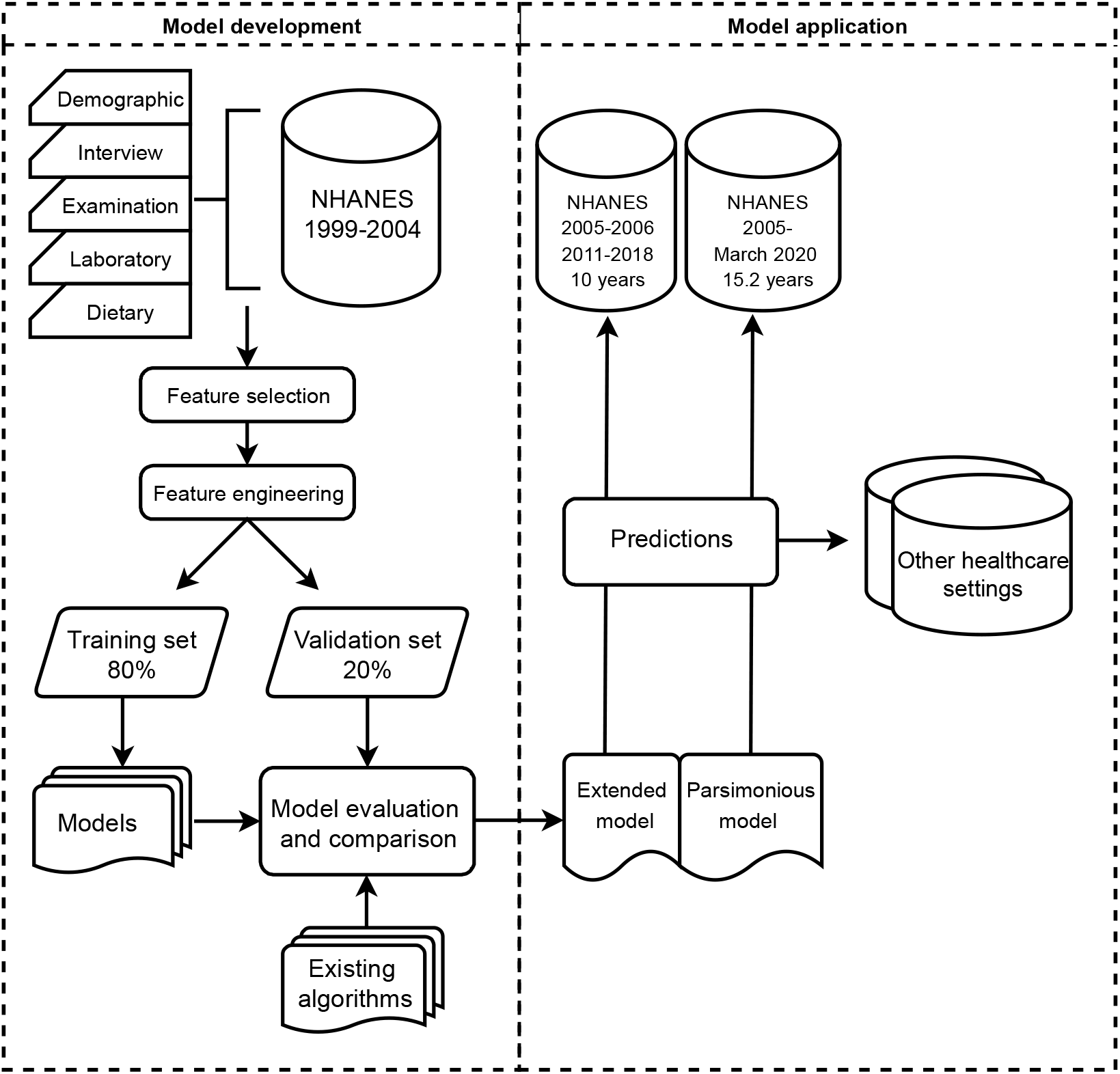
Flow chart visualizing the model development, evaluation, and prediction pipeline.

**Figure 2.**
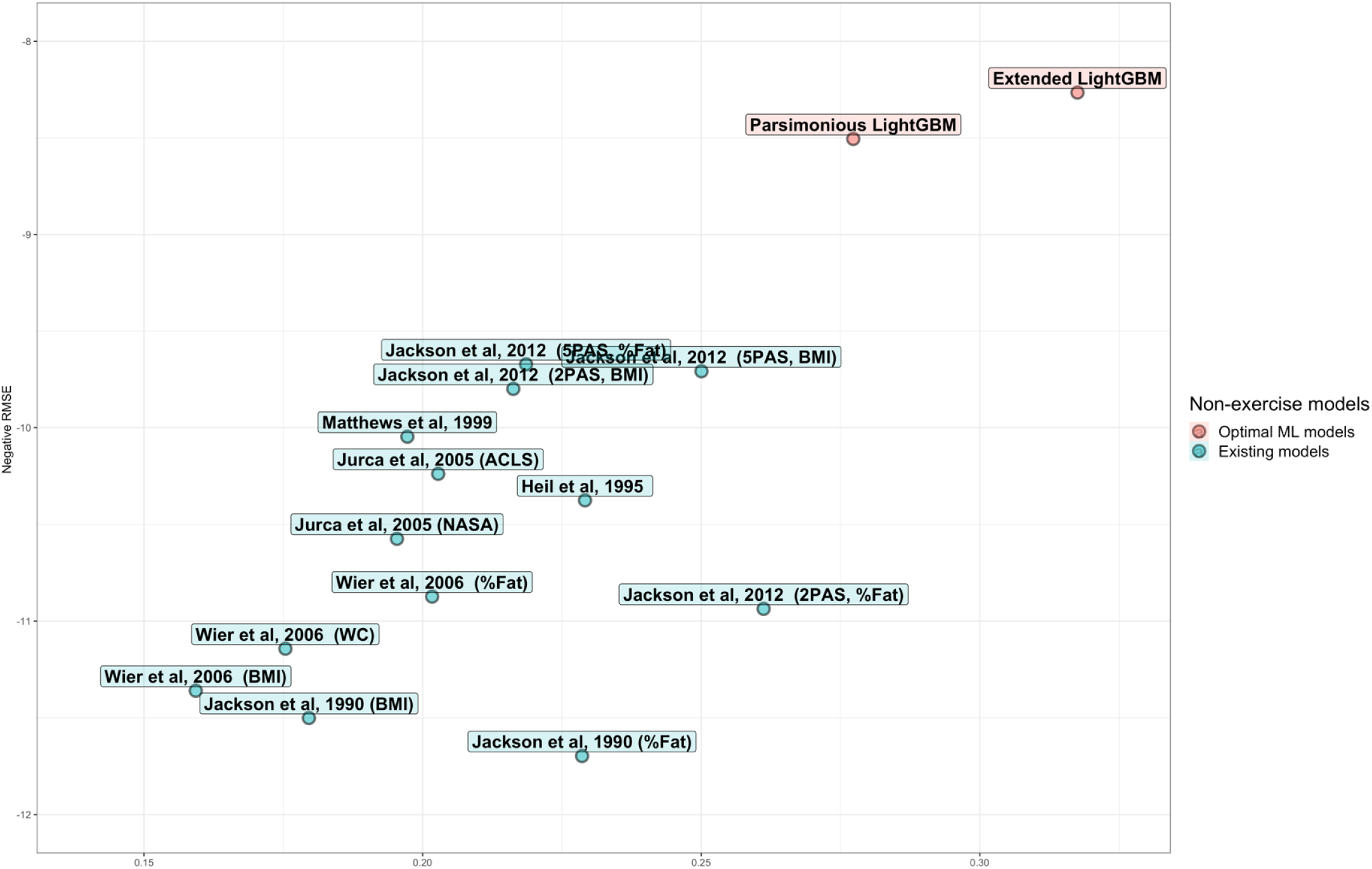
Visualization for model comparison, in which the y-axis represents the negative RMSE on the testing set, and the x-axis indicates the coefficient of determination on the testing set. Each point represented a specific model. Newly developed ML models and the existing non-exercise models were differentiated by color. The better models tended to show up in the upper righter corner of this figure. (Abbreviation, LightGBM: Light Gradient Boosting Machine, 5PAS: 5 level-Physical activity score, 2PAS: 2 level-Physical activity score, %Fat: total percent fat, WC: Waist Circumference, BMI: Body Mass Index, NASA/JSC: National Aeronautics and Space Administration/Johnson Space Center, ACLS: Aerobics Center Longitudinal Study)

### Feature importance

We selected the top 16 relevant features for each optimal ML model based on the overall feature importance scores provided by SHAP. The SHAP analysis on the optimal parsimonious LightGBM model revealed that, in order of importance, gender, waist circumference, the 60s resting pulse rate, weekly total moderate-intensity activity time, BMI, age, height, race and ethnicity, family poverty income ratio, highest education level, daily alcohol intake, diastolic blood pressure weight, weight difference with last year, systolic blood pressure, and smoking status are the key predictors of VO_2_max. The extended LightGBM model’s SHAP values additionally identified the averaged leg percent fat, averaged arm percent fat, trunk percent fat, calcium, total cholesterol, creatinine, leg bone mineral density (BMD), potassium, BUN (blood urea nitrogen), and trunk lean as important features (**Figure 3**). Specifically, considering only the continuous features, the percent fat of leg, arm, and trunk, the 60s resting pulse rate, age, calcium, poverty income ratio, total cholesterol, diastolic blood pressure, creatinine, waist circumference, BMI, weight, and systolic blood pressure are negatively associated with the predicted VO_2_max.

**Figure 3.**
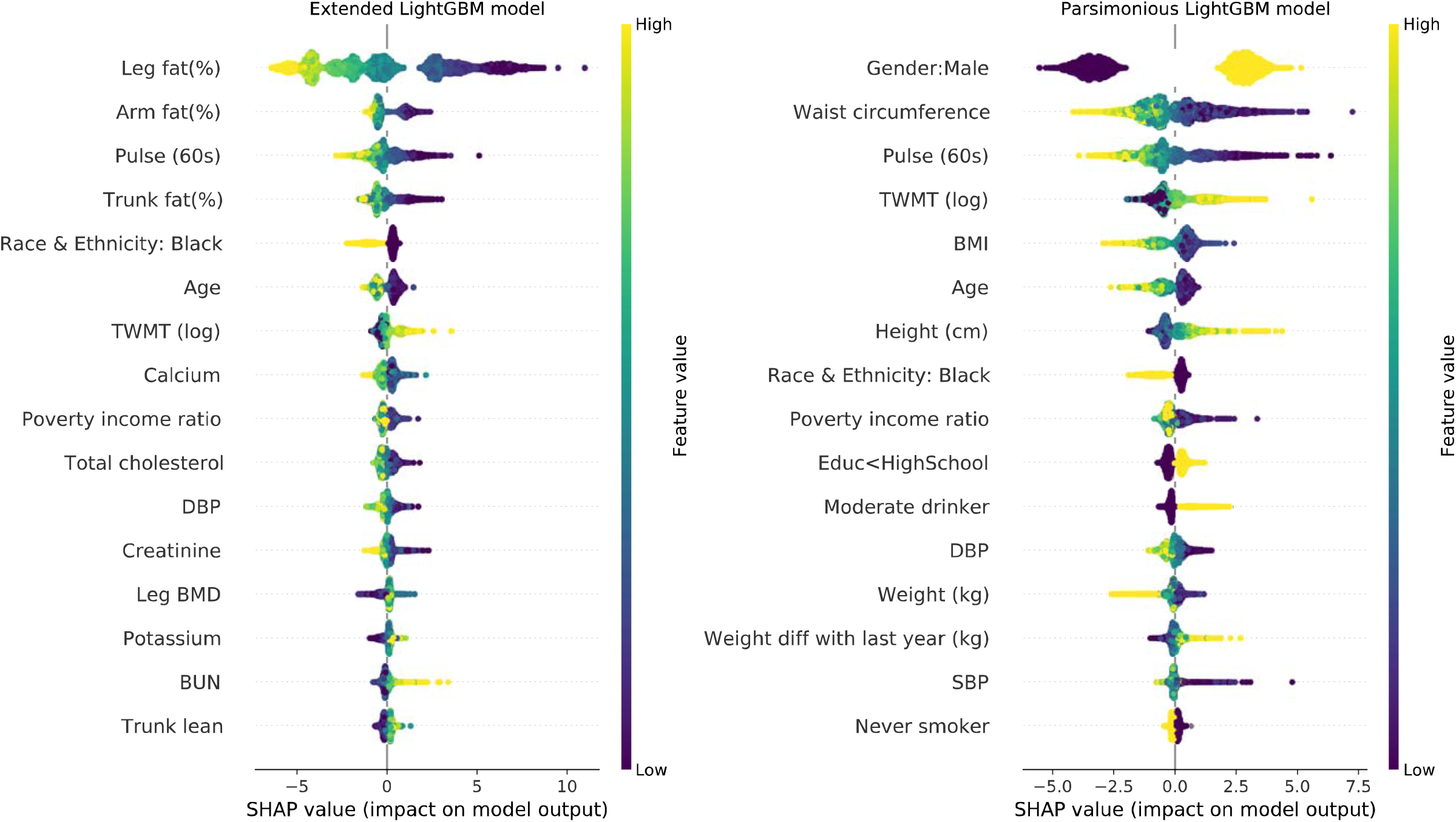
SHAP feature importance for both the parsimonious and the extended VO_2_max prediction models. To provide as much insight into the contribution of each feature, we used a SHAP summary plot, in which the y-axis represents the feature (in descending order of overall importance), and the x-axis indicates the change/effect each feature exerted on the prediction for each participant (as represented by a dot). The gradient color denoted the magnitude of the original value for that feature. More negative SHAP values (left on the x-axis) indicated stronger negative impacts on predicted VO_2_max.

The higher values those features are, the lower values those features would make the predicted VO_2_max become. As for the weekly total moderate-intensity activity time, BUN, trunk lean, height, and weight loss in one year, they are positively associated with the predicted VO_2_max (**Figure 3, Supplemental Figure S9, S10**).

### Prediction for NHANES

The extended and the parsimonious model predictions had similar distributions to the exercise test measurements from 1999-2004 (**Supplemental Figure S8**). Sex-specific nationally representative estimates of the mean VO2max were reported in **Figure 4**. Different types of estimates were relatively consistent across NHANES cycles, and no significant trends or changes in VO2max at the national scale were found.

**Figure 4.**
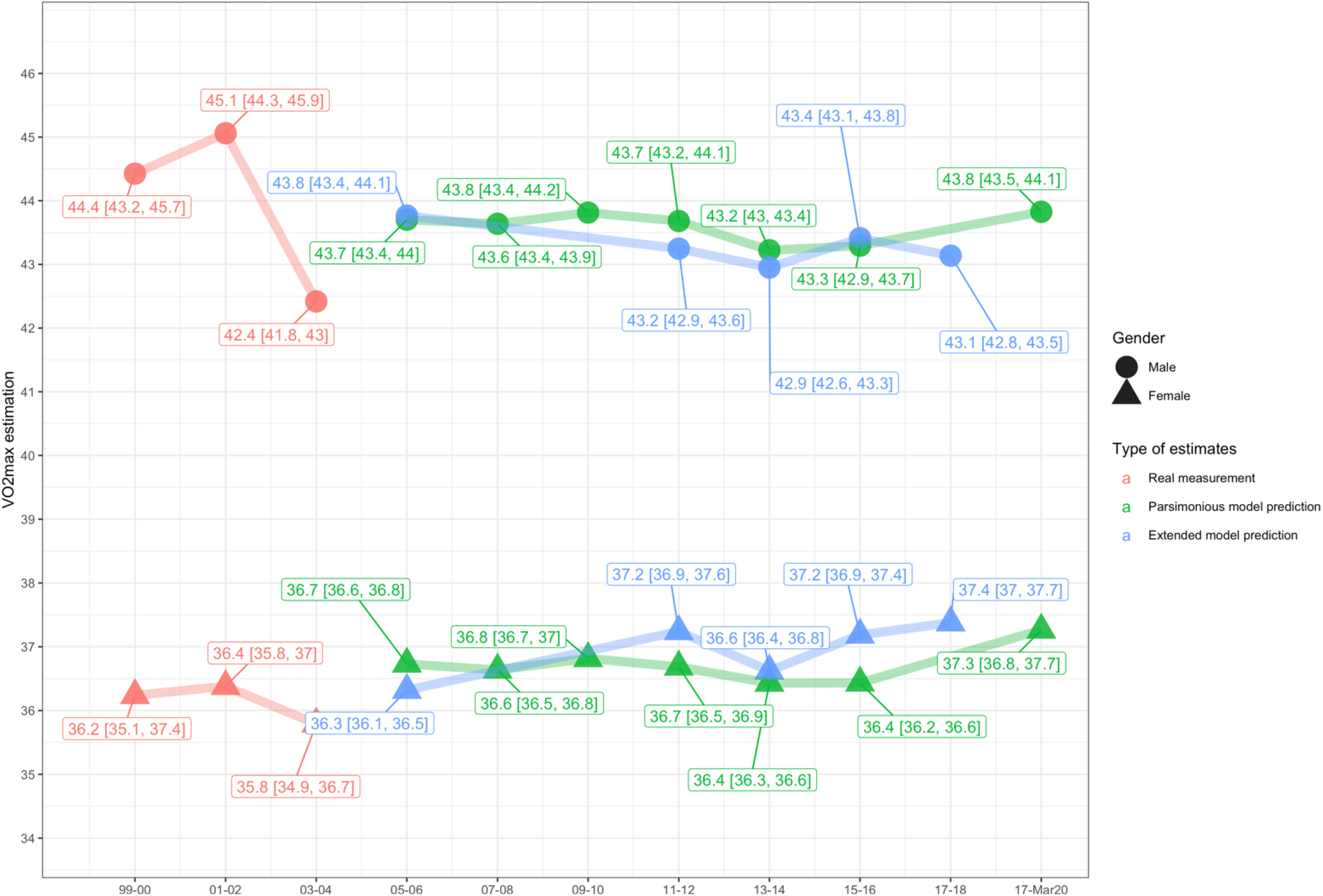
Nationally representative estimate of mean VO_2_max by gender over the eligible NHANES subjects across different cycles. Figure 4 was colored by the type of VO_2_max outcome, the type depended on whether the VO_2_max was measured in NHANES, predicted by the optimal parsimonious model or the extended model. The different shapes differentiated gender.

## DISCUSSION

The ML models developed in this study outperformed existing non-exercise algorithms for predicting VO_2_max. Such models can be applied to NHANES cycles that lacked the CRF component to make nationally representative estimates of cardiorespiratory fitness. In addition, this model may be useful in clinical situations where there is a need for an estimate of the VO_2_max in the absence of an exercise test.

The algorithm developed in this study advances the field in several ways. First, we used the NHANES data, a national data source, to develop our models. Therefore, models built on NHANES data have excellent generalizability, which allows us to provide a nationally representative estimation. Most previous studies depended on local data or were biased towards specific subgroups with poor external validity. For instance, the NASA study population was only representative of the NASA/Johnson Space Center workforce,^19,20^ the ACLS study consisted mostly of white participants who were of higher SES.^8,9^

Second, we used a more comprehensive set of variables to develop the models and applied state-of-the-art ML algorithms to address non-linear associations and interactions among the predictors. The comprehensive set of variables explored in this study came from both the NHANES interviews and the physical examination sections, which included demographic, socioeconomic, and health-related questions, medical and physiological measurements, and laboratory tests administered by highly trained medical personnel. By using ML algorithms, our models had stronger predictive power, and they were more efficient to train. Our models significantly outperformed the previous methods, which had a limited number of predictors and used mostly linear models.

Third, we applied the improved models to generate nationally representative estimates of CRF, thereby providing new insights into cardiorespiratory health in the US population. NHANES CRF data were measured in 1999-2004 only, such that national data on fitness in the U.S. has been lacking ever since 2005. We demonstrated, in NHANES, the feasibility of predicting the discontinued measurement of VO_2_max with the model generated from cycles that had VO_2_max (in the CRF component) recorded. The predicted outcomes showed consistency with the real measurements in the form of nationally representative estimates, they could be considered reliable approximates for those cycles that lacked VO_2_max data.

Our findings have important clinical and public health implications. First, using accessible national survey data and ML algorithms, we demonstrated the feasibility to develop reliable prediction models of CRF and comprehensively evaluate the CRF status of the U.S population. These models can be calibrated and generalized to other populations to predict CRF. Moreover, we provide a useful pipeline for researchers and policymakers to leverage existing national data to predict important population health variables that were not consistently available, either missing in earlier cycles or discontinued to be recorded in later periods. Second, the nationally representative estimates of the predicted VO_2_max, along with the ones from the real measurements, provided insights into the cardiorespiratory fitness status of the US population over the recent 20 years. These results will enable a better understanding of overall cardiovascular health and offer a reference for the formation of relevant public health guidelines and policies. Another implication was the newly discovered factors that correlated to CRF. Besides the commonly acknowledged predictors, we found family income, BUN, blood pressure, cholesterol, leg bone mineral density, trunk lean, and creatinine also important in the CRF prediction. Regarding fat percent for different parts of the body (leg, trunk, arm), we found the averaged leg percent fat had the highest relevance with CRF. The disparity in fat percent of different body parts as risk factors is worthy of future research.

Our study has several limitations. First, the extended models built in this study were cross-sectional. Although they furnished reliable estimates of CRF at the population level, they could not assess changes in CRF. The effect of time was neglected when developing the models in this study, leaving time interactions unevaluated. Second, the available CRF outcomes in NHANES were derived from the submaximal exercise test, which is not the optimal way of measuring CRF. In a submaximal exercise test, rather than being directly measured, VO_2_max is calculated with the assumption that the relation between heart rate and oxygen consumption is linear during exercise. Therefore, the models and the corresponding predictions may be subject to the errors or violations introduced by the indirect property of the submaximal exercise test. Third, although the models built in this study were expected to have better generalizability, they were still limited by the exclusion criterion and selected variables in NHANES. The exclusion criterion NHANES adopted narrowed down the study population to relatively healthy younger-age (16-49) people who qualified for the submaximal exercise test, which constrained the use of the model in the elderly population. Also, the various kinds of NHANES variables used in this study might not be available in other healthcare settings. Thus, the actual feasibility of the use of our models outside the scope of NHANES remains to be determined by external validation, and it requires extra attention to the study population either for applying the models or interpreting the results from the models. Finally, we only considered CRF as the continuous outcome in this study while clinical measurements are more useful as prognostic indicators when a specified level of the parameter being measured identifies a threshold of increased risk for adverse health outcomes. Currently, there is no complete consensus on a level of CRF that classifies an asymptomatic individual as high risk, nor is there agreement as to what level of CRF is sufficient in the context of health and disease prevention. We did not add information on this important topic, but such insight can be gained by examining the association between the VO_2_max predicted by the model and other significant health outcomes in future NHANES studies. In all, additional work is needed to assess the applicability of this approach in primary care and other settings, to verify the validity of non-exercise estimates of CRF as predictors of health outcomes, and to establish a target level of CRF for primary prevention, placing CRF levels into clinical relevance like guidelines for blood pressure or body mass index.

In summary, we presented the ML models built from NHANES data to predict VO_2_max. We demonstrated the viability of using these models to be a proxy for direct measurements via exercise testing. They can make more accurate predictions and reliable national representative estimates for CRF than the previously published models.

## Data Availability

All data produced are available online at https://wwwn.cdc.gov/nchs/nhanes/Default.aspx

https://wwwn.cdc.gov/nchs/nhanes/Default.aspx

## Non-standard Abbreviations and Acronyms

CRF: Cardiorespiratory fitness
VO_2_max: Maximal oxygen uptake
CPX: Cardiopulmonary exercise testing
ML: Machine learning
NHANES: National Health and Nutrition Examination Survey
STROBE: Strengthening the Reporting of Observational Studies in Epidemiology
COVID-19: coronavirus disease 2019
MEC: Mobile Examination Center
KNN: K-Nearest Neighbors
LASSO: Least Absolute Shrinkage and Selection Operator
SVR: Support Vector Regression
RF: Random Forest
GBDT: Gradient Boosting decision tree
XGBoost: Extreme Gradient Boosting
LightGBM: Light Gradient Boosting Machine
SHAP: Shapley additive explanation

## Funding

None.

## Disclosures

In the past three years, Harlan Krumholz received expenses and/or personal fees from UnitedHealth, Element Science, Aetna, Reality Labs, Tesseract/4Catalyst, F-Prime, the Siegfried and Jensen Law Firm, Arnold and Porter Law Firm, and Martin/Baughman Law Firm. He is a co-founder of Refactor Health and HugoHealth, and is associated with contracts, through Yale New Haven Hospital, from the Centers for Medicare & Medicaid Services and through Yale University from Johnson & Johnson. Bobak Mortazavi received expenses and/or personal fees from HugoHealth, as a consultant. Dr. Khera receives support from the National Heart, Lung, and Blood Institute of the National Institutes of Health under award, 1K23HL153775, and is a founder of Evidence2Health, a precision health and digital health analytics platform. The other co-authors report no potential competing interests.

## Supplemental Material

Tables S1–S4

Figure S5-S9

References 37-43, 48-49

## Notes

### Funding Statement

This study did not receive any funding

### Author Declarations

The study used (or will use) ONLY openly available human data that were originally located at:https://wwwn.cdc.gov/nchs/nhanes/Default.aspx

## REFERENCES

1. Caspersen CJ, Powell KE, Christenson GM. Physical activity, exercise, and physical fitness: definitions and distinctions for health-related research. Public health reports. 1985;100:126.

2. Ross R, Blair SN, Arena R, Church TS, Després J-P, Franklin BA, Haskell WL, Kaminsky LA, Levine BD, Lavie CJ. Importance of assessing cardiorespiratory fitness in clinical practice: a case for fitness as a clinical vital sign: a scientific statement from the American Heart Association. Circulation. 2016;134:e653–e699.

3. Lee DC, Sui X, Artero EG, Lee IM, Church TS, McAuley PA, Stanford FC, Kohl HW, 3rd, Blair SN. Long-term effects of changes in cardiorespiratory fitness and body mass index on all-cause and cardiovascular disease mortality in men: the Aerobics Center Longitudinal Study. Circulation. 2011;124:2483–2490. doi: 10.1161/CIRCULATIONAHA.111.038422

4. Lee CD, Blair SN, Jackson AS. Cardiorespiratory fitness, body composition, and all-cause and cardiovascular disease mortality in men. Am J Clin Nutr. 1999;69:373–380. doi: 10.1093/ajcn/69.3.373

5. Lakka TA, Venalainen JM, Rauramaa R, Salonen R, Tuomilehto J, Salonen JT. Relation of leisure-time physical activity and cardiorespiratory fitness to the risk of acute myocardial infarction. N Engl J Med. 1994;330:1549–1554. doi: 10.1056/NEJM199406023302201

6. Kodama S, Saito K, Tanaka S, Maki M, Yachi Y, Asumi M, Sugawara A, Totsuka K, Shimano H, Ohashi Y, et al. Cardiorespiratory fitness as a quantitative predictor of all-cause mortality and cardiovascular events in healthy men and women: a meta-analysis. JAMA. 2009;301:2024–2035. doi: 10.1001/jama.2009.681

7. Ekelund LG, Haskell WL, Johnson JL, Whaley FS, Criqui MH, Sheps DS. Physical fitness as a predictor of cardiovascular mortality in asymptomatic North American men. The Lipid Research Clinics Mortality Follow-up Study. N Engl J Med. 1988;319:1379–1384. doi: 10.1056/NEJM198811243192104

8. Blair SN, Kohl HW, 3rd, Paffenbarger RS, Jr., Clark DG, Cooper KH, Gibbons LW. Physical fitness and all-cause mortality. A prospective study of healthy men and women. JAMA. 1989;262:2395–2401. doi: 10.1001/jama.262.17.2395

9. Blair SN, Kampert JB, Kohl HW, 3rd, Barlow CE, Macera CA, Paffenbarger RS, Jr., Gibbons LW. Influences of cardiorespiratory fitness and other precursors on cardiovascular disease and all-cause mortality in men and women. JAMA. 1996;276:205–210.

10. Lee D-c, Artero EG, Sui X, Blair SN. Review: Mortality trends in the general population: the importance of cardiorespiratory fitness. Journal of Psychopharmacology. 2010;24:27–35. doi: 10.1177/1359786810382057

11. Medicine ACoS. ACSM’s guidelines for exercise testing and prescription. Lippincott Williams & Wilkins; 2013.

12. Mitchell JH, Sproule BJ, Chapman CB. The physiological meaning of the maximal oxygen intake test. The Journal of clinical investigation. 1958;37:538–547.

13. Taylor HL, Buskirk E, Henschel A. Maximal oxygen intake as an objective measure of cardio-respiratory performance. Journal of applied physiology. 1955;8:73–80.

14. Foster C, Jackson AS, Pollock ML, Taylor MM, Hare J, Sennett SM, Rod JL, Sarwar M, Schmidt DH. Generalized equations for predicting functional capacity from treadmill performance. American heart journal. 1984;107:1229–1234.

15. Astrand PO, Ryhming I. A nomogram for calculation of aerobic capacity (physical fitness) from pulse rate during sub-maximal work. J Appl Physiol. 1954;7:218–221. doi: 10.1152/jappl.1954.7.2.218

16. Jackson AS, Blair SN, Mahar MT, Wier LT, Ross RM, Stuteville JE. Prediction of functional aerobic capacity without exercise testing. Med Sci Sports Exerc. 1990;22:863–870. doi: 10.1249/00005768-199012000-00021

17. Survey ADNF. A report on activity patterns and fitness levels. In: Sports Council and Health Education Authority London; 1992.

18. Heil DP, Freedson PS, Ahlquist LE, Price J, Rippe JM. Nonexercise regression models to estimate peak oxygen consumption. Med Sci Sports Exerc. 1995;27:599–606.

19. JackSon AS, Beard EF, Wier LT, Ross RM, Stuteville JE, Blair SN. Changes in aerobic power of men, ages 25-70 yr. Medicine and science in sports and exercise. 1995;27:113–120.

20. Jackson AS, Wier LT, Ayers GW, Beard EF, Stuteville JE, Blair SN. Changes in aerobic power of women, ages 20-64 yr. Medicine and science in sports and exercise. 1996;28:884–891.

21. Williford HN, Scharff-Olson M, Wang N, Blessing DL, Smith FH, Duey WJ. Cross-validation of non-exercise predictions of VO2peak in women. Med Sci Sports Exerc. 1996;28:926–930. doi: 10.1097/00005768-199607000-00022

22. George JD, Stone WJ, Burkett L. Non-exercise VO2max estimation for physically active college students. Medicine and science in sports and exercise. 1997;29:415–423.

23. Matthews CE, Heil DP, Freedson PS, Pastides H. Classification of cardiorespiratory fitness without exercise testing. Med Sci Sports Exerc. 1999;31:486–493. doi: 10.1097/00005768-199903000-00019

24. Jurca R, Jackson AS, LaMonte MJ, Morrow JR, Jr., Blair SN, Wareham NJ, Haskell WL, van Mechelen W, Church TS, Jakicic JM, et al. Assessing cardiorespiratory fitness without performing exercise testing. Am J Prev Med. 2005;29:185–193. doi: 10.1016/j.amepre.2005.06.004

25. Wier LT, Jackson AS, Ayers GW, Arenare B. Nonexercise models for estimating VO2max with waist girth, percent fat, or BMI. Med Sci Sports Exerc. 2006;38:555–561. doi: 10.1249/01.mss.0000193561.64152

26. Mailey EL, White SM, Wojcicki TR, Szabo AN, Kramer AF, McAuley E. Construct validation of a non-exercise measure of cardiorespiratory fitness in older adults. BMC Public Health. 2010;10:59. doi: 10.1186/1471-2458-10-59

27. Nes BM, Janszky I, Vatten LJ, Nilsen TI, Aspenes ST, Wisloff U. Estimating V.O 2peak from a nonexercise prediction model: the HUNT Study, Norway. Med Sci Sports Exerc. 2011;43:2024–2030. doi: 10.1249/MSS.0b013e31821d3f6f

28. Jackson AS, Sui X, O’Connor DP, Church TS, Lee D-c, Artero EG, Blair SN. Longitudinal cardiorespiratory fitness algorithms for clinical settings. American journal of preventive medicine. 2012;43:512–519.

29. Molnar C. Interpretable machine learning. Lulu. com; 2020.

30. Obermeyer Z, Emanuel EJ. Predicting the future—big data, machine learning, and clinical medicine. The New England journal of medicine. 2016;375:1216.

31. Chen L. Overview of clinical prediction models. Annals of translational medicine. 2020;8.

32. Herrin J, Abraham NS, Yao X, Noseworthy PA, Inselman J, Shah ND, Ngufor C. Comparative effectiveness of machine learning approaches for predicting gastrointestinal bleeds in patients receiving antithrombotic treatment. JAMA network open. 2021;4:e2110703–e2110703.

33. Von Elm E, Altman DG, Egger M, Pocock SJ, Gøtzsche PC, Vandenbroucke JP. The Strengthening the Reporting of Observational Studies in Epidemiology (STROBE) statement: guidelines for reporting observational studies. Bulletin of the World Health Organization. 2007;85:867–872.

34. Stierman B, Afful J, Carroll MD, Chen T-C, Davy O, Fink S, Fryar CD, Gu Q, Hales CM, Hughes JP. National Health and Nutrition Examination Survey 2017–March 2020 Prepandemic Data Files Development of Files and Prevalence Estimates for Selected Health Outcomes. 2021.

35. Hyattsville. The NHANES Cardiovascular Fitness Procedure Manual. In: Statistics NCfH, ed.; 2004.

36. Maranhão Neto GdA, Lourenço PMC, Farinatti PdTV. Prediction of aerobic fitness without stress testing and applicability to epidemiological studies: a systematic review. Cadernos de saude publica. 2004;20:48–56.

37. Fix E, Hodges JL. Discriminatory Analysis - Nonparametric Discrimination - Consistency Properties. International Statistical Review. 1989;57:238–247. doi: Doi 10.2307/1403797

38. Tibshirani R. Regression shrinkage and selection via the Lasso. Journal of the Royal Statistical Society Series B-Methodological. 1996;58:267–288. doi: DOI 10.1111/j.2517-6161.1996.tb02080.x

39. Cortes C, Vapnik V. Support-vector networks. Machine Learning. 1995;20:273–297. doi: 10.1007/bf00994018

40. Breiman L. Random Forests. Machine Learning. 2001;45:5–32. doi: 10.1023/a:1010933404324

41. Friedman JH. Greedy function approximation: a gradient boosting machine. Annals of statistics. 2001:1189–1232.

42. Chen T, Guestrin C. Xgboost: A scalable tree boosting system. Paper/Poster presented at: Proceedings of the 22nd acm sigkdd international conference on knowledge discovery and data mining; 2016;

43. Ke G, Meng Q, Finley T, Wang T, Chen W, Ma W, Ye Q, Liu T-Y. Lightgbm: A highly efficient gradient boosting decision tree. Advances in neural information processing systems. 2017;30.

44. Lundberg SM, Erion G, Chen H, DeGrave A, Prutkin JM, Nair B, Katz R, Himmelfarb J, Bansal N, Lee S-I. From local explanations to global understanding with explainable AI for trees. Nature machine intelligence. 2020;2:56–67.

45. Lundberg SM, Lee S-I. A unified approach to interpreting model predictions. Advances in neural information processing systems. 2017;30.

46. Shapley L. Notes on n-person games VII. cores of convex games. In: RAND CORP SANTA MONICA CALIF; 1965.

47. Van Buuren S, Groothuis-Oudshoorn K. mice: Multivariate imputation by chained equations in R. Journal of statistical software. 2011;45:1–67.

48. Hyattsville. The nhanes cardiovascular fitness procedure manual. 2004

49. Piercy KL, Troiano RP, Ballard RM, Carlson SA, Fulton JE, Galuska DA, et al. The physical activity guidelines for americans. Jama. 2018;320:2020–2028

