## Supplemental material for "Non-exercise Machine Learning Models for Maximal Oxygen Uptake Prediction in National Population Surveys"

### Table S1. Exclusion Criteria Classification for the NAHNES Cardiovascular Fitness Component (1999-2004) ^48^

| **Age** | - Age <16 years old, or age >=50 years old |
| --- | --- |
| **Physical functioning limitations** | - Difficulties in walking for a quarter mile - Difficulties in walking up 10 steps without resting - Difficulties in walking from one room to another on the same level - Difficulties in standing up from an armless straight chair - Having back or neck problems - Having fractures or injuries in bone or joint - Having health problem that requires the use of special equipment, such as a cane or a wheelchair - Having a bone or joint problem that could be made worse by walking - Lose balance due to dizziness on a regular basis - Lose consciousness on a regular basis - Blind or with very poor eyesight - Diabetes affected eyes or had retinopathy - Having developmental problems - Having amputations of legs or feet other than toes - Weight exceeded equipment limitation (>350 lb) - Other specified physical limitations |
| **Cardiovascular conditions/symptoms** | - Had been diagnosed with congestive heart failure - Had been diagnosed with coronary heart disease - Had been diagnosed with angina - Had been diagnosed with myocardial infarction - Had been diagnosed with stroke - Self-reported heart problems - Self-reported stroke problems - Having a pacemaker or automatic defibrillator - Doctor had instructed to do only physical activity recommended by a doctor because of a heart condition - Feeling chest pain during physical activity - Had chest pain when not doing physical activity - Resting heart rate ≥ 100 beats/min - Resting systolic blood pressure ≥ 180 mmHg - Resting diastolic blood pressure ≥ 100 mmHg - Irregular heartbeats: 3 or more dropped beats in 30 seconds |
| **Lung/breathing conditions/symptoms** | - Having to stop for breath when walking at own pace on level ground - Having to stop for breath after walking about 100 yards or after a few minutes on level ground - Having been awakened by trouble breathing or shortness of breath - Having to sleep on 2 or more pillows to help breathe - Self-reported lung or breathing problems - Had been diagnosed with emphysema |
| **Asthma symptoms** | - Had 12 or more attacks of wheezing or whistling during the past 12 months |
| **Medication exclusions** | - Anti Arrhythmics - Amiodarone (Cordarone) - Bretylium (Bretylol) - Cardioquin (Quinidine, Quinalan, Quinidex, Quinaglute, Quinora) - Disopyramide (Norpace) - Encainide (Enkaid) - Flecainide (Tambocor) - Lidocaine (Xylocaine, Xylocard) - Metoprolol Succinate (Toprol-XL) - Mexiletine (Mexitil) - Moricizine (Ethmozine) - Procainamide (Pronestyl, Procan SR) - Propafenone (Rhythmol) - Sotolol (Betapace) - Tocainide (Tonocard) - Beta Blockers - Acebutolol (Monitan, Sectral) - Atenolol (Tenormin) - Betaxolol (Kerlone) - Bisoprolol (Zebeta) - Carteolol (Cartrol) - Carvedilol (Coreg) - Esmolol (Brevibloc) - Labetalol (Normodyne) - Metoprolol tartrate (Betaloc, Lopressor) - Nadolol (Corgard) - Oxprenolol (Trasicor, Slow-Trasicor) - Penbutolol (Levatol) - Pindolol (Visken) - Propranolol (Detensol, Inderal) - Sotolol (Betapace, Sotocor) - Timolol (Blocadren) - Beta Blockers/Diuretic Combinations - Atenolol and Chlorthalidone (Tenorectic) - Bisoprolol and Hydrochlorothiazide (Ziac) - Metoprolol and Hydrochlorothiazide (Lopressor HCT) - Nadolol and Bendroflumethiazide (Corzide) - Propranolol and hydrochlorothiazide (Inderide) - Timolol and Hydrochlorothiazide (Timolide) - Calcium Channel-Blockers - Amlodipine (Norvasc) - Amlodipine and Atorvastatin (Caduet) - Amlodipine and Valsartan (Exforge) - Bepridil (Vascor) - Diltiazem (Cardizem, Ciatia, Dilacor, Diltia, Tiazac) - Felodipine (Plendil) - Isradipine (DynaCirc) - Mibefradil (Posicor) - Nicardipine (Cardene) - Nifedipine (Adalat, Nifedical, Procardia) - Nimodipine (Nimotop) - Nisoldipine (Sular) - Verapamil (Covera, Verelan, Calan, Isoptin) - CNS Stimulant - Ephedrine (Ma Huang) - Digitalis - Digoxin (Lanoxin) - Eye Drops/ Beta Blockers - Betaxolol Ophthalmic (Betoptic) - Timolol Ophthalmic (Timoptic) - Levobunolol Ophthalmic (betagan) - Metipranolol Ophthalmic (OptiPranolol) - Nitrates and Nitroglycerin - Erythrityl tetranitrate (Cardiwell) - Isosorbide dinitrate (Diltrate, Isordil, Nitrolin) - Isosorbide mononitrate (Monoket, Ismo) - Nitroglycerin, sustained release (Nitrong, Nitrocine, Nitroglyn) - Nitroglycerin, topical (Deponit, Minitran, Nitro-Dur, Nitrodisc, Nitrek, Nitrol, Nitro-Bid, Transderm Nitro, Nitroderm) - Nitroglycerin, translingual (Duotrate, Nitrostat, Nitrolingual spray, Isonate) - Nitroglycerin, transmucosal (Nitrogard) - Pentaerythritol tetranitrate (Cardilate, Peritrate) - Ephedra Based Weight Loss Medication - Dymetadrine Xtreme - Extreme Ripped Force - Metabolife - Metabolift - Phentermine - Pro-Ripped Ephedra - Ripped Fuel - Stacker - Ultra Ripped |
| **Other specified reasons** | - Had been hospitalized for specified reasons in the past 3 months (see the CV Fitness Procedures Manual, Appendix D for details) - Doctor recommended not to participate in sports or other activities due to a health condition - Other safety concerns specified by the participant - Other safety concerns identified by MEC physician or staff |

### Table S2. Exclusion Criteria the NAHNES Cardiovascular Fitness prediction (2005-March 2020)

- Age <16 years old, or age >=50 years old
- Pregnant
- Difficulties in walking for a quarter of a mile
- Difficulties in walking up 10 steps without resting
- Blind or with very poor eyesight
- Had been diagnosed with congestive heart failure
- Had been diagnosed with coronary heart disease
- Had been diagnosed with angina
- Had been diagnosed with myocardial infarction
- Had been diagnosed with stroke
- Self-reported heart problems
- Self-reported stroke problems

**Table S3. Extended Machine learning models**

| Algorithms | Description | Reason |
| --- | --- | --- |
| **K-Nearest neighbors (KNN)** | KNN^37^ makes regression predictions for an observation based on the outcome of its k closest neighbors which defined by a metric measuring the distance between the observation and other cases from the data sample from the feature space. | KNN is a simple but effective non-parametric supervised learning method. It does not make any presumptions on the input dataset. It is easy to be implemented and a good choice for a benchmark. |
| **Least absolute shrinkage and selection operator (LASSO)** | LASSO^38^ is a kind of linear model with L1-regularization. Lasso achieves regularization and feature selection by forcing the sum of the absolute value of the regression coefficients to be less than a fixed value, which forces certain coefficients to zero, excluding them from impacting prediction. | LASSO performs both regularization and variable selection, it provides good generalizability and interpretability. It can address the issue of collinearity in linear models. |
| **Support vector regression (SVR)** | Support vector regression (SVR)^39^ is derived from the idea of Support vector machine. It is characterized by using kernels, sparse solution, control of the margin and the number of support vectors | SVR has been proven to be a very effective tool in real-value function estimation. It is robust to outliers. Besides, the size of the data in our study is suitable for the use of SVR. |
| **Random Forest**  **(RF)** | Random forest^40^ is an ensemble method that operates by constructing a multitude of decision trees through a bagging method. Each tree generated from the randomly and independently sampled observations and their features. For regression tasks, the mean or average prediction of the individual trees is returned. Random forest often does a good job on prediction task with well controlled variance | Random forest is handy to utilize. It achieves low bias with acceptable variance. It is robust to outliers and it has great generalizability. Each tree trained in the random forest algorithm is interpretable. |
| **Extreme Gradient Boosting (XGBoost**) | XGBoost^43^ is an implementation of gradient boosted decision trees. Gradient boosting decision tree (GBDT) is an ensemble method based on growing decision trees successively using gradient descent to minimize a loss function. It connects the stagewise additive expansions and the steepest-descent minimization because one new base model is added at a time and existing base models in the ensemble model are frozen and left unchanged. | XGBoost generally performs well in the prediction performance for tasks in various domains. It can deal with non-linear relationships and features’ interaction without extra processing. It is robust to outliers and has satisfying generalizability. It is also efficient to train and relatively convenient to use. It has a built-in capability to handle missing values |
| **Light Gradient Boosting Machine (LightGBM)** | LightGBM^43^ is another implementation of gradient decision trees. It grows decision tree leaf-wise. | LightGBM basically inherits all the strengths of XGBoost, but LightGBM runs is much faster than XGBoost and LightGBM is more suitable for larger dataset. |

**Table S4.** Definition of predictor variables in NHANES.

|  | **Predictor variable** | **Section** | **Data type** | **Ascertainment in NHANES** | **Definition of variables used in analysis** | **Missing rate (%)** |
| --- | --- | --- | --- | --- | --- | --- |
| **Parsimonious**  **Model** | Age | Demographic | Continuous | Best age in years of the sample person at the time of HH screening. Individuals 85 and over are top coded at 85 years of age.  Related NHANES variable:   - RIDAGEYR (1999-2020) | Age in years of the sample person at time of HH screening. | 0 |
|  | Gender | Demographic | Binary | Gender of the sample person  Related NHANES variable:   - RIAGENDR (1999-2020) | Gender is classified as:   - Male - Female | 0 |
|  | Race and ethnicity | Demographic | Categorical | Reported race and ethnicity information.  Related NHANES variable:   - RIDRETH1 (1999-2020) | Race and ethnicity is classified:   - Non-Hispanic White - Non-Hispanic Black - Hispanic - Others | 0 |
|  | Poverty income ratio | Demographic | Continuous | Poverty income ratio (PIR) - a ratio of family income to poverty threshold. The PIR was calculated by dividing family income by the poverty guidelines, specific to family size, as well as the appropriate year and state. PIR values greater than or equal to 5.00 are top coded as 5.0.  Related NHANES variable:   - INDFMPIR (1999-2020) | The ratio of family income to poverty threshold | 7.9 |
|  | Family income | Demographic | Binary | Index for the ratio of family income to poverty.  Related NHANES variable:   - INDFMPIR (1999-2020) | Based on the poverty income ratio relative to the federal poverty limit from the Census Bureau, family income is categorized as   - High/middle income [≥2] - Low-income [<2] | 7.9 |
|  | Marital status | Demographic | Binary | Marital Status. Marital status data were collected for sample persons 14 years of age and older  Related NHANES variable:   - DMDMARTL (1999-2020) | Marital status is classified as:   - married/ living with partners - others (widowed, divorced, or separated; and never married). | 2.1 |
|  | Highest education level | Demographic | Categorical | Highest grade or level of school completed, or the highest degree received?  Related NHANES variable:   - DMDEDUC2 (1999-2020) - DMDEDUC3 (1999-2020) | Highest education level is classified as:   - Less than high school - High school - Greater than high school | 0.0 |
|  | Health insurance | Questionnaire | Binary | In-person interview: Are you covered by health insurance or some other kind of health care plan? [Include health insurance obtained through employment or purchased directly as well as government programs like Medicare and Medicaid that provide medical care or help pay medical bills.]  Related NHANES variable:   - HID010 (1999-2004) - HIQ011 (2005-2020) | Health insurance is classified:   - 1: if participant answered “Yes” - 0: if participant answered “No” | 1.7 |
|  | Smoking status | Questionnaire | Categorical | In-person interview:   - Ever smoke cigarettes in entire life (SMQ620, SMQ620) - Smoked at least 100 cigarettes in life (SMQ020) - Do you now smoke cigarettes? (SMQ040) - Used tobacco/nicotine last 5 days?   (SMD680, SMQ680, SMQ681)  Related NHANES variable:   - SMQ020 (1999-2020) - SMQ040 (1999-2020) - SMQ620 (1999-2010) - SMQ621 (2011-2020) - SMD680 (1999-2001) - SMQ680 (2001-2012) - SMQ681 (2013-2020) | Smoking status is defined as:   - Never smoker: defined as individuals who stated they have never smoked cigarettes in entire life or smoked less than 100 cigarettes in life. - Former smokers: defined as individuals who stated they had smoked cigarettes, or smoked at least 100 cigarettes in life, but in the meantime stated they do not smoke now, or did not use tobacco/nicotine last 5 days - Current smokers: defined as individuals who stated they smoke now, or used tobacco/nicotine last 5 days | 1.9 |
|  | Total weekly moderate-intensity activity time (TWMT) | Questionnaire | Continuous | **1999-2005:**   - Over the past 30 days, did any vigorous activities for at least 10 minutes that caused heavy sweating, or large increases in breathing or heart rate? Some examples are running, lap swimming, aerobics classes or fast bicycling (PAD200) - Over the past 30 days, did any moderate activities for at least 10 minutes that cause only light sweating or a slight to moderate increase in breathing or heart rate? Some examples are brisk walking, bicycling for pleasure, golf, and dancing. (PAD320) - Reported intensity level of activity (PADLEVEL) - Frequency of the activity (PADTIMES) - On average about how long for the activity each time? (PADTIMES)   **2007-2020:**   - Exclude the work and transportation activities, do any vigorous-intensity sports, fitness, or recreational activities that cause large increases in breathing or heart rate like running or basketball for at least 10 minutes continuously? (PAQ650) - Do any moderate-intensity sports, fitness, or recreational activities that cause a small increase in breathing or heart rate such as brisk walking, bicycling, swimming, or golf for at least 10 minutes continuously? (PAD665) - In a typical week, on how many days do vigorous-intensity sports, fitness, or recreational activities? (PAQ655) - In a typical week, on how many days do moderate-intensity sports, fitness, or recreational activities? (PAD670) - How much time spent on doing vigorous-intensity sports, fitness, or recreational activities on a typical day? (PAD660) - How much time spent on doing moderate-intensity sports, fitness, or recreational activities on a typical day? (PAD675) | The time of vigorous-intensity activity is equivalent to twice the time of moderate-intensity activity. Moderate-intensity aerobic activity time can be approximated by 2 times vigorous-intensity activity time.^49^  Total weekly moderate-intensity activity time is calculated as, in a typical week, total moderate-intensity activity time, which adds up the original moderate-intensity activity time and the equivalent moderate-intensity activity time from vigorous activity. | 0.0 |
|  | Physical activity | Questionnaire | Categorical | Based on the NHANES variables previously defined total weekly moderate-intensity activity time (TWMT) and TWMT itself. | Physical activity is classified as:   - Recommended: 150 min or more of TWMT - Insufficient: TWMT between 10-149 min   Inactive: no participation or less than 10 min of TWMT | 0.0 |
|  | Weekly Walk/bicycle times (days) | Questionnaire | Continuous | - Over the past 30 days, walked or bicycled as part of getting to and from work, or school, or to do errands? (PAD020) - Over the past 30 days, how often walk or bicycle as part of getting to and from work, or school, or to do errands PROBE: How many times per day, per week, or per month do these activities? (PAQ050Q) - Walk or use a bicycle for at least 10 minutes continuously to get to and from places? (PAQ635) - In a typical week, on how many days walk or bicycle for at least 10 minutes continuously to get to and from places? (PAD640)   Related NHANES variable:   - PAD020 (1999-2006) - PAD640 (2007-2018) | - Using the frequency and time to generate the weekly Walk/bicycle times (in days), ranging from 0 to 7 days. | 0.0 |
|  | Try to lose weight last year | Questionnaire | Binary | - During the past 12 months, tried to lose weight? (WHQ070)   Related NHANES variable:   - WHQ070 (1999-2020) | Coded as:   - 1: if participant answered “Yes”   0: if participant answered “No” | 9.0 |
|  | Weight difference current with last year | Questionnaire | Continuous | - Self-reported weight - 1 year ago (pounds) (WHD050) - Current self-reported weight (pounds) (WHD020)   Related NHANES variable:   - WHD020 (1999-2020) - WHD050 (1999-2020) | - Current self-reported weight minus the self-reported weight 1 year ago. | 2.4 |
|  | Exercised to lose weight / keep from gaining weight | Questionnaire | Binary | - Did you exercise to weight? (WHD080D) - What did you do to keep from gaining weight? (WHD100D)   Related NHANES variable:   - WHD080D (1999-2020)   WHD100D (1999-2020) | Coded as:   - 1: if participant answered “Yes” or provided the code for exercise.   0: if participant answered “No” or did not response | 2.7 |
|  | History of hypertension | Questionnaire | Binary | - Has a doctor or other health professional ever told you that you had hypertension, also called high blood pressure?   Related NHANES variable:   - BPQ020 (1999-2020) | Coded as:   - 1: if participant answered “Yes” - 0: if participant answered “No” or did not response. | 2.3 |
|  | SBP | Examination | Continuous | - Systolic: Blood pressure mm Hg   Related NHANES variable:  BPXSYx (1999-2020, x denotes the x^th^ reading) | - Averaged systolic blood pressure over all readings. | 0.0 |
|  | DBP | Examination | Continuous | - Diastolic: Blood pressure mm Hg   Related NHANES variable:   - BPXDIx (1999-2020, x denotes the x^th^ reading) | Averaged diastolic blood pressure over all readings. | 0.0 |
|  | BMI | Examination | Continuous | - Body Mass Index (kg/m**2)   Related NHANES variable:   - BMXBMI (1999-2020) | Body Mass Index (kg/m**2) | 0.4 |
|  | Waist circumference | Examination | Continuous | - Waist Circumference (cm)   Related NHANES variable:   - BMXWAIST (1999-2020) | Waist Circumference (cm) | 0.8 |
|  | Pulse rate (60s) | Examination | Continuous | - 60-seccond pulse   Related NHANES variable:   - BMXWAIST (1999-2020) | 60-second pulse | 0 |
|  | Weight (kg) | Examination | Continuous | Weight (kg)  Related NHANES variable:   - BMXWAIST (1999-2020) | Weight (kg) | 0.4 |
|  | Height (cm) | Examination | Continuous | Standing height (cm)  Related NHANES variable:   - BMXHT (1999-2020) | Standing height (cm) | 0.4 |
| Extended model  (Features added additionally on top of the parsimonious model) | Arm fat (%) | Examination | Continuous | Averaged Arm Percent Fat  Related NHANES variable:   - DXDRAPF   (1999-2005, 2011-2018)   - DXDLAPF   (1999-2005, 2011-2018) | Averaged Arm Percent Fat =  $\frac{DXDRAPF+DXDLAPF}{2}$ | 3.2 |
|  | Arm BMD | Examination | Continuous | Averaged Arm Body Mineral Density (grams/cm^2)  Related NHANES variable:   - DXXRABMD   (1999-2005, 2011-2018)   - DXXLABMD   (1999-2005, 2011-2018) | Averaged Arm Body Mineral Density (grams/cm^2) =  $\frac{DXXRABMD+DXXLABMD}{2}$ | 3.2 |
|  | Arm lean | Examination | Continuous | Averaged Arm Lean excl Bone Mineral Content (grams)  Related NHANES variable:   - DXDLALE   (1999-2005, 2011-2018)   - DXDRALE   (1999-2005, 2011-2018) | Averaged Arm Lean excl Bone Mineral Content (grams)=  $\frac{DXDLALE+DXDRALE}{2}$ | 3.2 |
|  | Leg fat (%) | Examination | Continuous | Averaged Leg Percent Fat  Related NHANES variable:   - DXDLLPF   (1999-2005, 2011-2018)   - DXDRLPF   (1999-2005, 2011-2018) | Averaged Leg Percent Fat =  $\frac{DXDLLPF+DXDRLPF}{2}$ | 3.2 |
|  | Leg BMD | Examination | Continuous | Averaged Leg Lean excl Bone Mineral Content (grams)  Related NHANES variable:   - DXXLLBMD   (1999-2005, 2011-2018)   - DXXRLBMD   (1999-2005, 2011-2018) | Averaged Arm Lean excl Bone Mineral Content (grams)=  $\frac{DXXLLBMD +DXXRLBMD}{2}$ | 3.2 |
|  | Leg lean | Examination | Continuous | Averaged Leg Lean excl Bone Mineral Content (grams)  Related NHANES variable:   - DXDLLLE   (1999-2005, 2011-2018)   - DXDRLLE   (1999-2005, 2011-2018) | Left Leg Lean excl Bone Mineral Content (grams)  $\frac{DXDLLLE+DXDRLLE}{2}$ | 3.2 |
|  | Trunk fat (%) | Examination | Continuous | Trunk Percent Fat  Related NHANES variable:   - DXDTRPF   (1999-2005, 2011-2018) | Trunk Percent Fat = DXDTRPF | 3.2 |
|  | Trunk BMD | Examination | Continuous | Trunk Bone BMD (g/cm^2)  Related NHANES variable:   - DXDTRBMD   (1999-2005, 2011-2018) | Trunk Bone BMD (g/cm^2) = DXDTRBMD | 3.2 |
|  | Trunk lean | Examination | Continuous | Trunk Lean excl Bone Mineral Content (grams)  Related NHANES variable:   - DXDTRLE   (1999-2005, 2011-2018) | Trunk Lean excl Bone Mineral Content (grams)= DXDTRLE | 3.2 |
|  | Alanine Aminotransferase  (ALT) | Laboratory | Continuous | Alanine aminotransferase ALT (U/L)  Related NHANES variable:  LBXSATSI (1999-2020) | Alanine aminotransferase ALT (U/L) | 5.0 |
|  | Aspartate Aminotransferase  (AST) | Laboratory | Continuous | Aspartate aminotransferase AST (U/L)  Related NHANES variable:  LBXSASSI (1999-2020) | Aspartate aminotransferase AST (U/L) | 5.0 |
|  | Blood Urea Nitrogen  (BUN) | Laboratory | Continuous | Blood urea nitrogen (mg/dL)  Related NHANES variable:  LBXSBU (1999-2020) | Blood urea nitrogen (mg/dL) | 4.9 |
|  | Glucose | Laboratory | Continuous | Glucose, refrigerated serum (mg/dL)  Related NHANES variable:  LBXSGL (1999-2020) | Glucose, refrigerated serum (mg/dL) | 4.9 |
|  | **Lactate Dehydrogenase**  (LDH) | Laboratory | Continuous | Lactate dehydrogenase (U/L)  Related NHANES variable:   - LBXSLDSI (1999-2000, 2003-2018)   LBDSLDSI (2001-2002) | Lactate dehydrogenase (U/L) | 5.0 |
|  | **Cholesterol** | Laboratory | Continuous | **Total Cholesterol, refrigerated serum (mg/dL)**  **Related NHANES variable:**   - **LBXSCH (1999-2020)** | **Total Cholesterol, refrigerated serum (mg/dL)** | **4.9** |
|  | Total protein | Laboratory | Continuous | **Total protein (g/dL)**  **Related NHANES variable:**   - **LBXSTP (1999-2020)** | **Total protein (g/dL)** | **5.0** |
|  | Potassium | Laboratory | Continuous | Potassium (mmol/L)  Related NHANES variable:   - LBXSKSI (1999-2020) | Potassium (mmol/L) | 5.0 |
|  | Sodium | Laboratory | Continuous | Sodium (mmol/L)  Related NHANES variable:   - LBXSNASI (1999-2020) | Sodium (mmol/L) | 4.9 |
|  | Chloride | Laboratory | Continuous | Chloride (mmol/L)  Related NHANES variable:   - LBXSCLSI (1999-2020) | Chloride (mmol/L) | 5.0 |
|  | Albuminuria | Laboratory | Continuous | Albumin, urine (mg/L)  Creatinine, urine (mg/dL)  Related NHANES variable:   - URXUMS (2005-2020) - URXUMA (1999-2004) - URXUMS (1999-2020) | Albuminuria =  $\frac{Albumin, urine (mg/L)}{Creatinine, urine (mg/dL)}*100$ | 10.4 |
|  | HBA1C | Laboratory | Continuous | Glycohemoglobin (%)  Related NHANES variable:  LBXGH (1999-2020) | Glycohemoglobin (%) | 3.9 |
|  | Creatinine | Laboratory | Continuous | Creatinine (mg/dL)  Related NHANES variable:   - LBXSCR (1999-2000, 2003-2020)   LBDSCR (2001-2002) | Creatinine (mg/dL) | 4.9 |
|  | Bilirubin | Laboratory | Continuous | Bilirubin, total (mg/dL)  Related NHANES variable:   - LBXSTB (1999-2000, 2003-2020)   LBDSTB (2001-2002) | Bilirubin, total (mg/dL) | 5.0 |
|  | Calcium | Laboratory | Continuous | Calcium, total (mg/dL)  Related NHANES variable:  LBXSCA (1999-2020) | Calcium, total (mg/dL) | 4.9 |
|  | Bicarbonate | Laboratory | Continuous | Bicarbonate (mmol/L)  Related NHANES variable:   - LBXSC3SI (1999-2020) | Bicarbonate (mmol/L) | 4.9 |
|  | Alcohol intake | Questionnaire | Categorical | In-person interview:   - In entire life, had at least 12 drinks of any type of alcoholic beverage. (ALQ110, ALQ111) - In any one year, had at least 12 drinks of any type of alcoholic beverage. (ALQ100, ALD100) - In the past 12 months, on those days that drank alcoholic beverages, on the average, how many drinks did you have (ALQ130) - For the Past 12 months, how often do you have alcohol drink (ALQ120U, ALQ120Q, ALQ121) - Use variables above calculate averaged daily drinks/alcohol intake   Related NHANES variable:   - ALQ100 (1999-2000) - ALD100 (2001-2002) - ALQ101 (2003-2016) - ALQ111 (2017-2018) - ALQ110 (1999-2016) - ALQ110 (1999-2016) - ALQ120 (1999-2016) - ALQ110 (1999-2016) - ALQ121 (2017-2018) | Alcohol intake status is categorized as:   - Never drinkers: defined as individuals who stated never had any kind of alcohol drink, or had less than 12 drinks of any type in entire life - Former drinkers: defined as individuals who stated they had had at least 12 drinks but did not drink for the past 12 months. - Light drinkers: defined as individuals who self-reported 0-0.5 average daily drinks. - Moderate drinkers: defined as individuals who self-reported (0.5-1.5 for women; 0.5-2.5 for men) average daily drinks. - Heavy drinkers: defined as individuals who self-reported (1.5+ for women; 2.5+ for men) average daily drinks. | 8.3 |

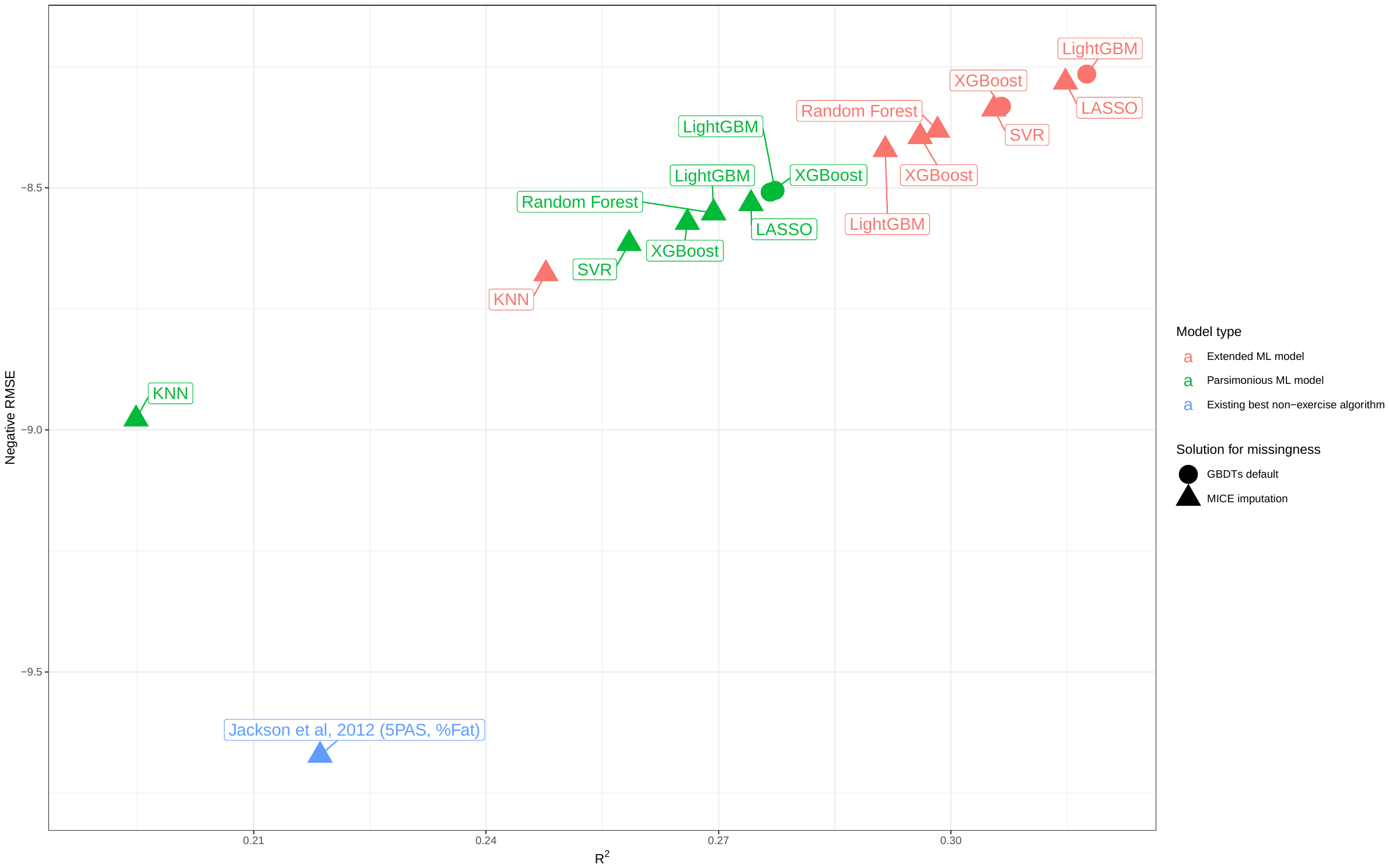

Figure S5 – Visualization for the newly developed ML model comparison, in which the y-axis represents the negative RMSE on the testing set, and the x-axis indicates the coefficient of determination on the training set. Each point represented a specific model. Different model types and solutions for missingness were noted by color and shape, respectively. Initially, for RMSE, a smaller value indicates better performance. Here we took the negative value of the RMSE, now a larger negative RMSE means better performance. Thus, the better models tended to show up in the upper righter corner of this figure. (Abbreviation, KNN: K-Nearest Neighbors, LASSO: Least absolute shrinkage and selection operator, SVR: Support vector regression, RF: Random Forest, XGBoost: Extreme Gradient Boosting, LightGBM: Light Gradient Boosting Machine)

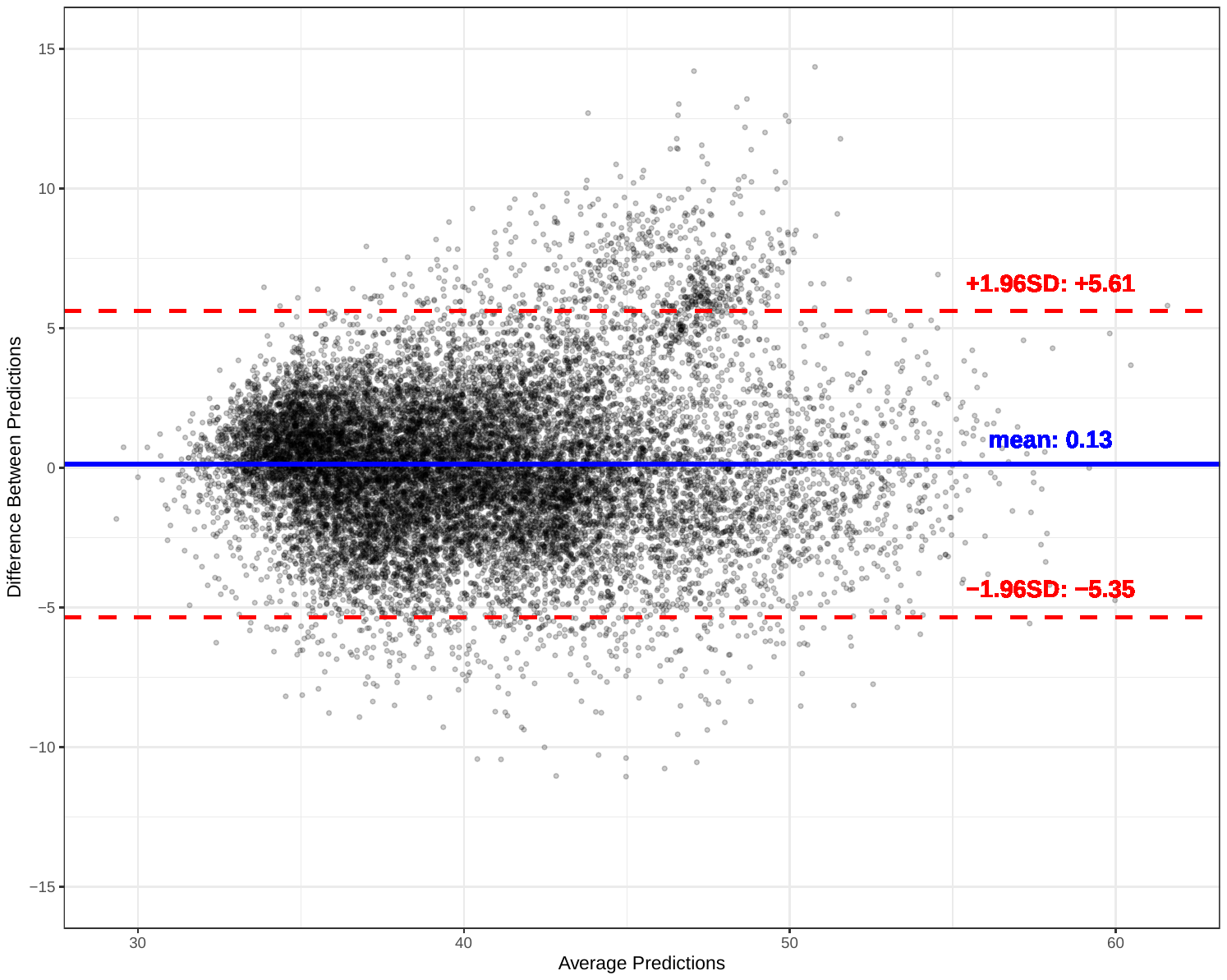

Figure S6 – Bland-Altman plot visualizing the agreement between the basic and the full LightGBM model.

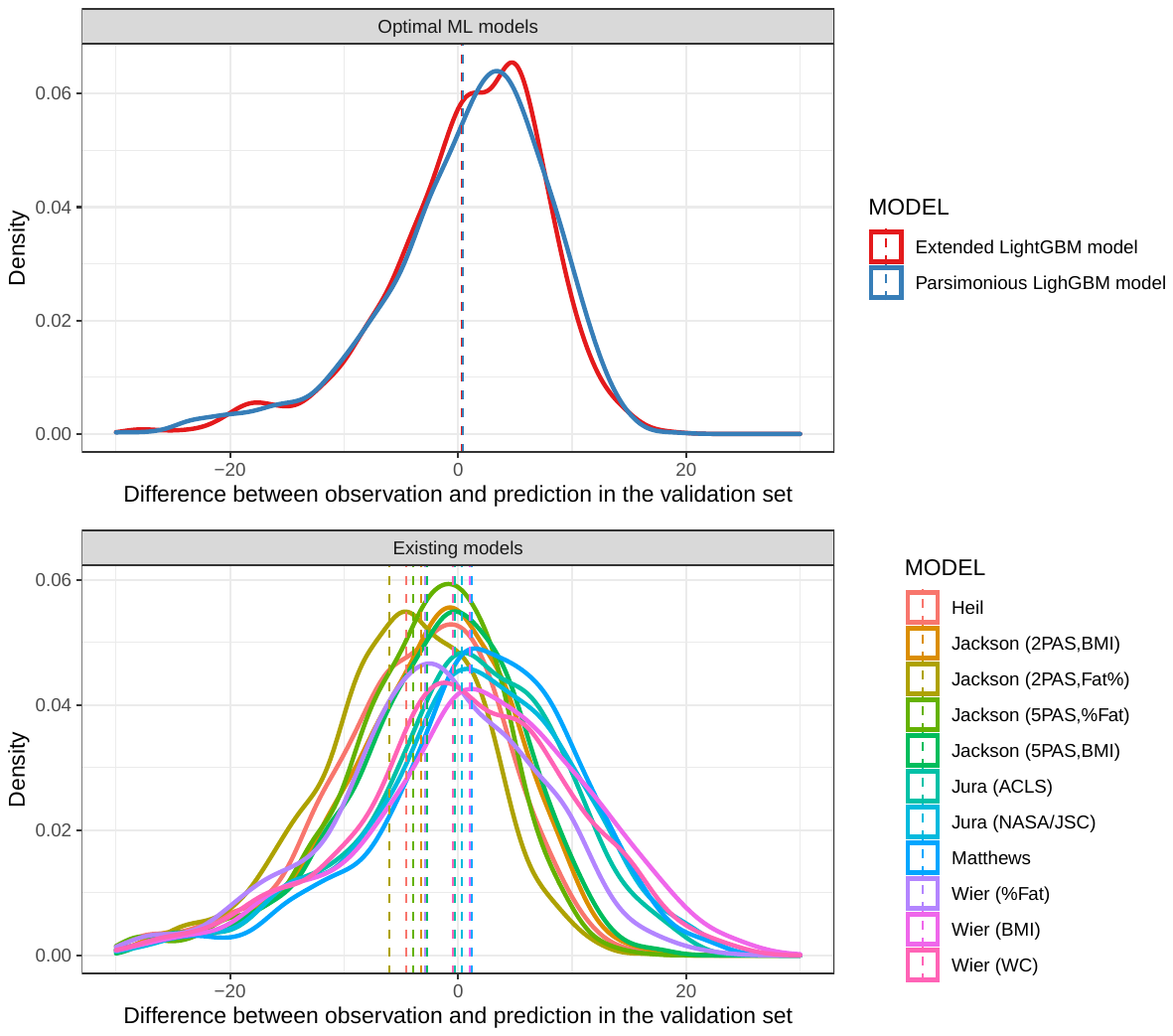

Figure S7 – Density curves describing the distributions of the differences between the observation and the prediction provided by the optimal ML models and existing non-exercise algorithms in the validation set.

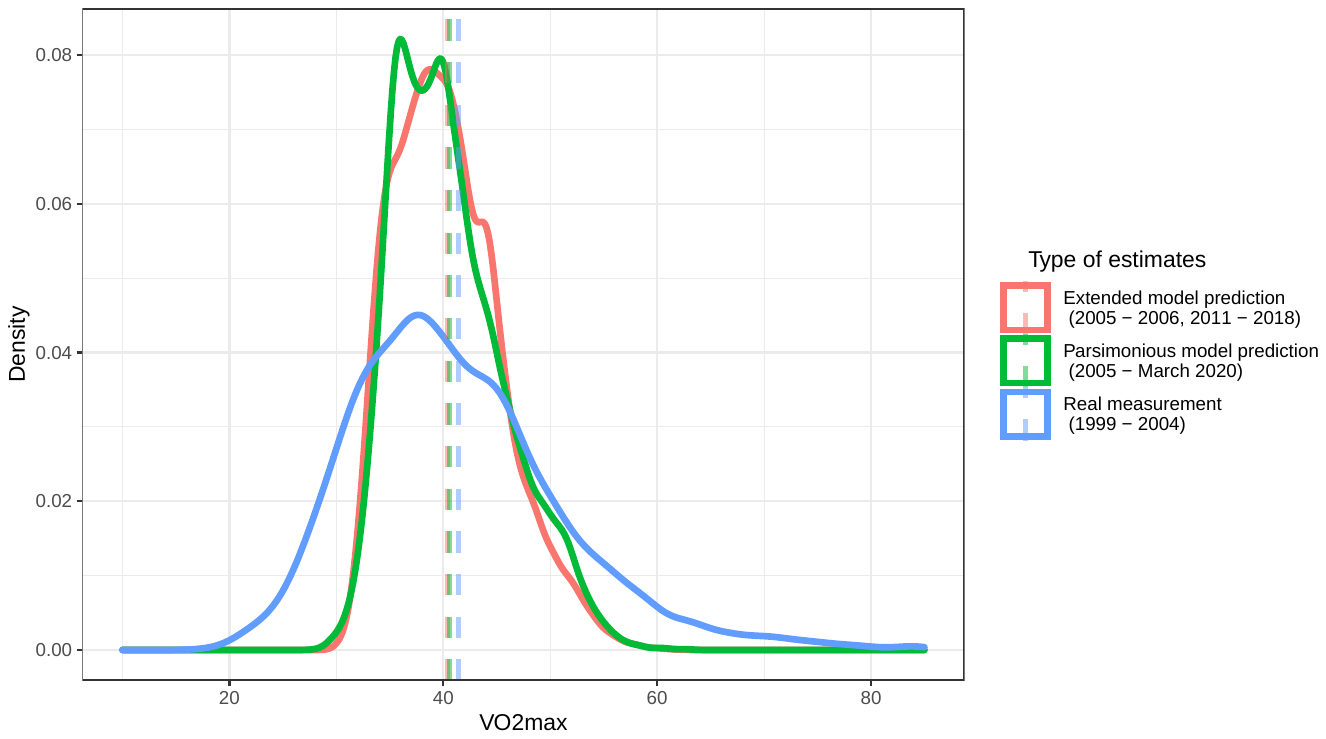

Figure S7 – Density curves describing the distributions of observation of measured VO2max in 1999-2004, and the predictions of VO2max from ML models after 2004.

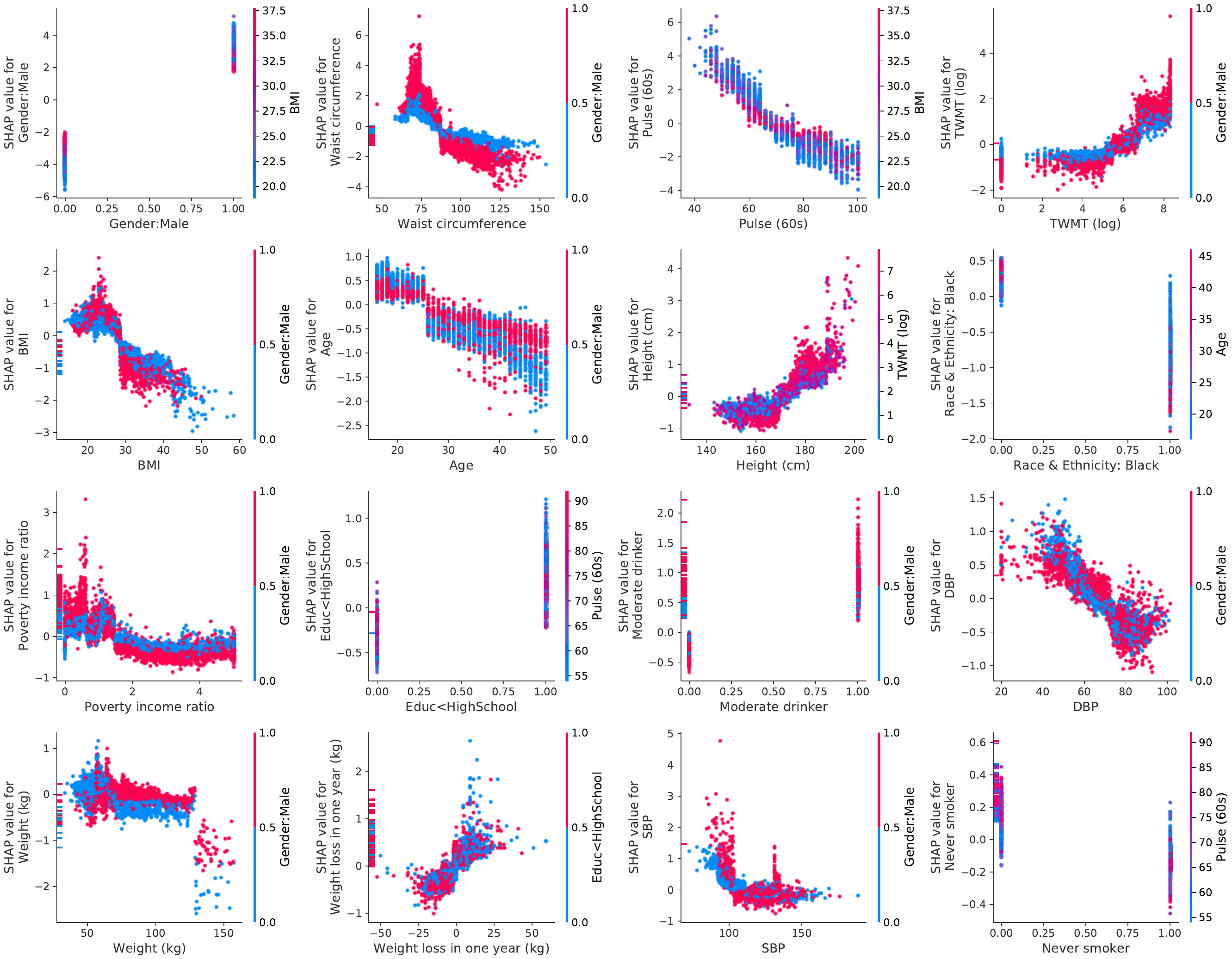

Figure S9– SHAP dependence plot for the parsimonious LightGBM, in which the x-axis is the value of the feature, and the y-axis is the SHAP value for that feature, which represents how much knowing that feature’s value changes the output of the model for the sample’s prediction. Each dot is a single prediction from the dataset, The color corresponds to a second feature that may have an interaction effect with the feature plotted, by default this second feature is chosen automatically by SHAP algorithms. (Abbreviation, TWMT (log): Total Weekly Moderate-intensity activity Time (in log-transformation), SBP: Systolic Blood Pressure, DBP: Diastolic Blood Pressure, BMI: Body Mass Index)

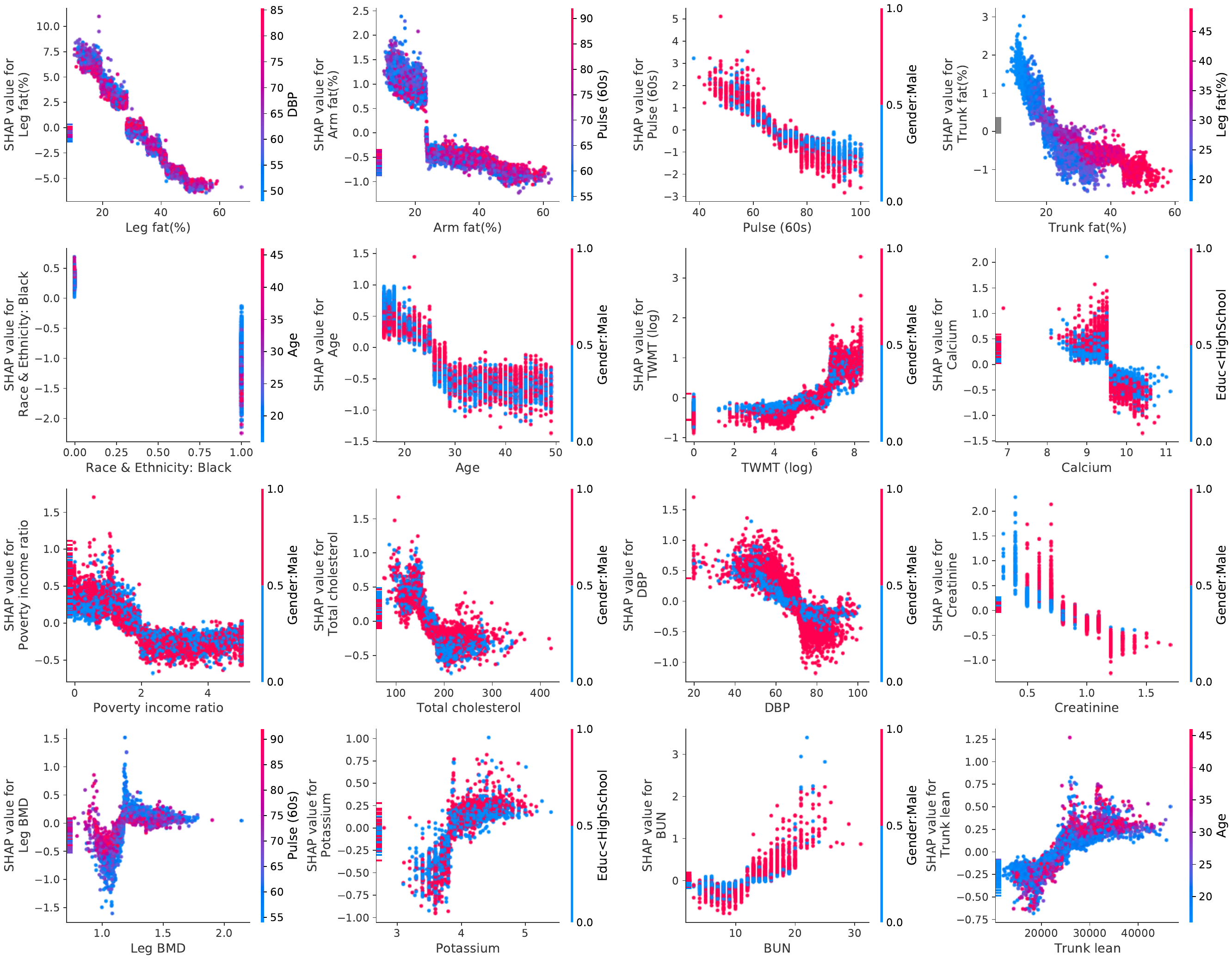

Figure S10 – SHAP dependence plot for the extended LightGBM, in which the x-axis is the value of the feature, and the y-axis is the SHAP value for that feature, which represents how much knowing that feature’s value changes the output of the model for the sample’s prediction. Each dot is a single prediction from the dataset, The color corresponds to a second feature that may have an interaction effect with the feature plotted, by default this second feature is chosen automatically by SHAP algorithms. (Abbreviation, TWMT (log): Total Weekly Moderate-intensity activity Time (in log-transformation), DBP: Diastolic Blood Pressure, BMD: Bone Mineral Density, BUN: blood urea nitrogen)
